# Co-existing mental and somatic conditions in Swedish children with the avoidant restrictive food intake disorder phenotype

**DOI:** 10.1101/2024.03.10.24304003

**Authors:** Marie-Louis Wronski, Ralf Kuja-Halkola, Elin Hedlund, Miriam I. Martini, Paul Lichtenstein, Sebastian Lundström, Henrik Larsson, Mark J. Taylor, Nadia Micali, Cynthia M. Bulik, Lisa Dinkler

## Abstract

**Background:** Avoidant restrictive food intake disorder (ARFID) is a feeding and eating disorder, characterized by limited variety and/or quantity of food intake impacting physical health and psychosocial functioning. Children with ARFID often present with a range of psychiatric and somatic symptoms, and therefore consult various pediatric subspecialties; large-scale studies mapping comorbidities are however lacking. To characterize health care needs of people with ARFID, we systematically investigated ARFID-related mental and somatic conditions in 616 children with ARFID and >30,000 children without ARFID.

**Methods:** In a Swedish twin cohort, we identified the ARFID phenotype in 6–12-year-old children based on parent-reports and register data. From >1,000 diagnostic ICD-codes, we specified mental and somatic conditions within/across ICD-chapters, number of distinct per-person diagnoses, and inpatient treatment days between birth and 18^th^ birthday (90 outcomes). Hazard ratios (HR) and incidence rate ratios (IRR) were calculated.

**Findings:** Relative risks of neurodevelopmental, gastrointestinal, endocrine/metabolic, respiratory, neurological, and allergic disorders were substantially increased in ARFID (e.g., autism HR[CI_95%_]=9.7[7.5–12.5], intellectual disability 10.3[7.6–13.9], gastroesophageal reflux disease 6.7[4.6–9.9], pituitary conditions 5.6[2.7–11.3], chronic lower respiratory diseases 4.9[2.4–10.1], epilepsy 5.8[4.1–8.2]). ARFID was not associated with elevated risks of autoimmune illnesses and obsessive-compulsive disorder. Children with ARFID had a significantly higher number of distinct mental diagnoses (IRR[CI_95%_]=4.7[4.0–5.4]) and longer duration of hospitalizations (IRR[CI_95%_]=5.5[1.7–17.6]) compared with children without ARFID. Children with ARFID were diagnosed earlier with a mental condition than children without ARFID. No sex-specific differences emerged.

**Interpretation:** This study yields the broadest and most detailed evidence of co-existing mental and somatic conditions in the largest sample of children with ARFID to date. Findings suggest a complex pattern of health needs in youth with ARFID, underscoring the critical importance of attention to the illness across all pediatric specialties.

**Funding:** Fredrik and Ingrid Thurings Foundation, Mental Health Foundation.

**Research in context:** *Evidence before the study:* Avoidant restrictive food intake disorder (ARFID) is an eating and feeding disorder that often develops in childhood and that is associated with co-existing conditions such as anxiety; depression; and endocrine/metabolic, gastrointestinal, and immunological disorders. We systematically searched Embase, including Medline, and PubMed databases using the terms *(“avoidant restrictive food intake disorder” OR “ARFID”) AND (“comorbidity” OR ((“co-existing” OR “comorbid” OR “concurrent” OR “co-occurring”) AND (“concern” OR “condition” OR “disorder” OR “illness” OR “problem”)))* in title and abstract without language restrictions. Our search yielded 86 studies from 2013, when ARFID was first introduced as a diagnosis in DSM-5: most of these studies have been conducted in relatively small clinical samples, did not have a control group, or covered a limited range of mental and/or somatic conditions that often were reported as *concerns* rather than formal *diagnoses*. Further, we identified one systematic review (published in 2023) applying a more extensive search algorithm with similar terms, which reported that psychiatric comorbidity was common in ARFID, especially anxiety disorders (9–72%) and autism (8–55%). However, knowledge regarding medical treatment needs in ARFID is sparse; and potential effects of sex and age on co-existing conditions in ARFID throughout childhood and adolescence are unstudied, except for one study comparing comorbidities in 23 preschool vs. 28 school children with ARFID (published in 2023). The lack of large-scale studies on comorbidities in ARFID contributes to diagnostic misclassification and treatment delays, ultimately interfering with appropriate medical care.

*Added value of this study:* This cohort study, based on high-quality Swedish Twin Registry data, utilized the, to our knowledge, largest sample of children and adolescents with ARFID (n=616) *and* without ARFID (controls, n=30,179) to date. We applied a large-scale approach to study a broad range of mental and somatic diagnoses, received in both inpatient and outpatient settings, from birth to 18^th^ birthday (or until censored). In addition to confirming previous evidence of frequently co-occurring conditions such as attention deficit hyperactivity disorder, autism, and gastrointestinal disorders in a larger sample, we demonstrated that ARFID is associated with an increased risk of a wide variety of perinatal and congenital conditions (e.g., fetal growth retardation; perinatal jaundice and infections; circulatory, digestive, and nervous system malformations), allergic and respiratory conditions (e.g., acute and chronic lower and upper respiratory disorders), and psychiatric and neurodevelopmental disorders (e.g., tic disorders; conduct disorders; developmental disorders of motor function, speech and language, and scholastic skills). Furthermore, our study revealed greater treatment needs in children with ARFID compared with controls, indicated by higher number of distinct per-person diagnoses and longer duration of inpatient treatment due to any mental or somatic diagnosis (accumulated over time). Moreover, mental conditions were more likely to be diagnosed at an *earlier* age in children with AFRID relative to controls. However, this study did not yield relevant effects of sex assigned at birth on relative risk of any analyzed condition in ARFID vs. controls.

*Implications of all the available evidence:* Given the range and novelty of analyzed mental and somatic conditions, this study may generate hypotheses for future basic, epidemiological, and clinical research on the etiology, clinical presentation, and consequences of ARFID. Combined with previous evidence, we reveal the heterogenous and complex clinical presentations of the ARFID phenotype in childhood and adolescence. ARFID and its co-existing conditions require attention in the medical practice of multiple specialties (e.g., general pediatrics, pediatric endocrinology and gastroenterology, child and adolescent psychiatry, pediatric emergency care, family/internal medicine, and general practice) in order to develop multimodal diagnostic and treatment guidelines that improve treatment options for children and adolescents with ARFID.

## Introduction

Avoidant restrictive food intake disorder (ARFID) is a feeding and eating disorder with an estimated prevalence of 1–5% and a more even sex ratio than other eating disorders.^1,2^ ARFID is characterized by a severely limited range and/or quantity of food intake, with detrimental impact on weight/growth, nutritional status, and psychosocial functioning.^3^ In contrast to anorexia nervosa, ARFID is typically not triggered by body shape or weight concerns but rather by sensory aversion to smell, taste, or texture of food; low appetite or interest in food; and/or fear of aversive reactions to food intake such as choking or vomiting. Commonly emerging in childhood,^2^ the disorder is often persistent and associated with serious impairment.^4^

The identification and treatment of ARFID is complicated by frequently co-occurring mental and somatic health concerns, such as neurodevelopmental conditions (NDC) and gastrointestinal disorders^5^ like disorders of gut-brain interaction (DGBI)^6^ and esophagitis.^7^ This leads to complex clinical presentations, missed diagnosis, and delayed treatment.^7,8^ Commonly reported among youth with ARFID are elevated depressive (7–33%) and anxiety (9–72%) symptoms;^9,10^ endocrine alterations (e.g., thyroid and appetite-regulating hormone alterations);^11,12^ immunological conditions (e.g., asthma, food and drug allergies);^11^ and digestive problems (19–44%, e.g., acid reflux, delayed gastric emptying, irritable bowel syndrome).^5,7,9,11^ Further, frequent co-occurrence of ARFID and NDCs, particularly autism (8– 55% in ARFID) and attention deficit hyperactivity disorder (ADHD; 3-39% in ARFID), has been documented.^9,13^

There is a stark lack of research on comorbidities associated with ARFID, compared with other eating disorders.^14^ Most evidence on comorbidities in ARFID is limited to case reports and single-site studies conducted in relatively small or uncontrolled clinical samples.^7,8,10,11,15^ Reported health problems stem from different sources of varying data quality, including parent- or self-report medical history besides formal clinical diagnoses. Furthermore, effects of sex and age with respect to co-existing conditions in ARFID might pose important implications for treatment and prognosis, but are largely unknown.^15^ A deeper understanding of ARFID-related comorbidity could facilitate elucidating the etiology and sequelae of ARFID and inform diagnostic and therapeutic approaches.

Previously, we developed an ARFID phenotype in a sample of >30,000 Swedish twins via a diagnostic algorithm using parent-report and register data.^1^ Using the same sample (n=616 with ARFID), here we followed the children from birth until age 18 to systematically assess ARFID-related relative and absolute risks of a broad range of mental and somatic diagnoses. We also explored effects of sex and age. Moreover, we investigated treatment needs via number of per-person diagnoses and inpatient days. Building on previous evidence,^9,11,14^ we expected increased relative and absolute risks, higher number of per-person diagnoses, and longer hospitalization periods in children with ARFID vs. children without ARFID (henceforth referred to as *controls*) for most analyzed conditions.

## Methods

### Study design and participants

The study was conducted in accordance with the Helsinki Declaration. It was approved by the Regional Ethical Review Board in Stockholm, Sweden (Dnr 03-672, 2010/597-31/1, 2010/322-31/2). Informed assent was obtained from parents.

We retrieved relevant data from the Child and Adolescent Twin Study in Sweden (CATSS), which addressed all twins born in Sweden since July 1, 1992.^16^ CATSS is based on parent reports at twin age 9 years or 12 years (**Table S1**).^16^ Our study included 30,795 individual twins born between 1992 and 2008 (response rate approximately 69%), of whom 616 (2.00%) presented with the ARFID phenotype (**Table S1**).^1^

For defining the ARFID phenotype and study outcomes (i.e., mental/somatic conditions), we used data from the National Patient Register (NPR) until the end of 2016 (NPR contains all diagnostic and procedure codes from inpatient care since 1987 and ca. 80% of codes from specialized outpatient care since 2001; International Classification of Diseases (ICD)-9-coding used 1987–1996, ICD-10-coding since 1997)^17,18^ and the Prescribed Drug Register (PDR) until the end of 2017 (identified by their Anatomical Therapeutic Chemical [ATC] code).^19^ CATSS is linked to both NPR and PDR via anonymized identifiers.

### ARFID phenotype definition

The identification of the ARFID phenotype in this sample has been described extensively previously^1^: briefly, the composite measure includes CATSS parent-reports at twin age 9 or 12 years together with NPR diagnostic and treatment codes and PDR ATC-codes between twin age 6 and 12 years (**Figure 1**). Children were identified as having ARFID, if they met DSM-5 ARFID criterion A (avoidant/restrictive eating leading to clinical consequences, e.g., low weight, nutritional deficiencies, and psychosocial impairment) and DSM-5 ARFID criterion C (eating disturbance not attributable to anorexia nervosa, bulimia nervosa, or body image disturbance)^3^ between the age of 6 and 12 years. Data to assess DSM-5 ARFID Criterion B (eating disturbance not explained by lack of available food or associated culturally sanctioned practices)^3^ were not available. The goal of this study was to broadly map co-occurring conditions in ARFID. In epidemiological contexts, it is difficult to evaluate whether specific co-occurring conditions are cause, comorbidity, or consequence of avoidant/restrictive eating. Thus, we did *not* exclude ARFID based on co-occurring mental or somatic conditions that could potentially explain avoidant/restrictive eating (DSM-5 ARFID criterion D).^3^

**Figure 1.**
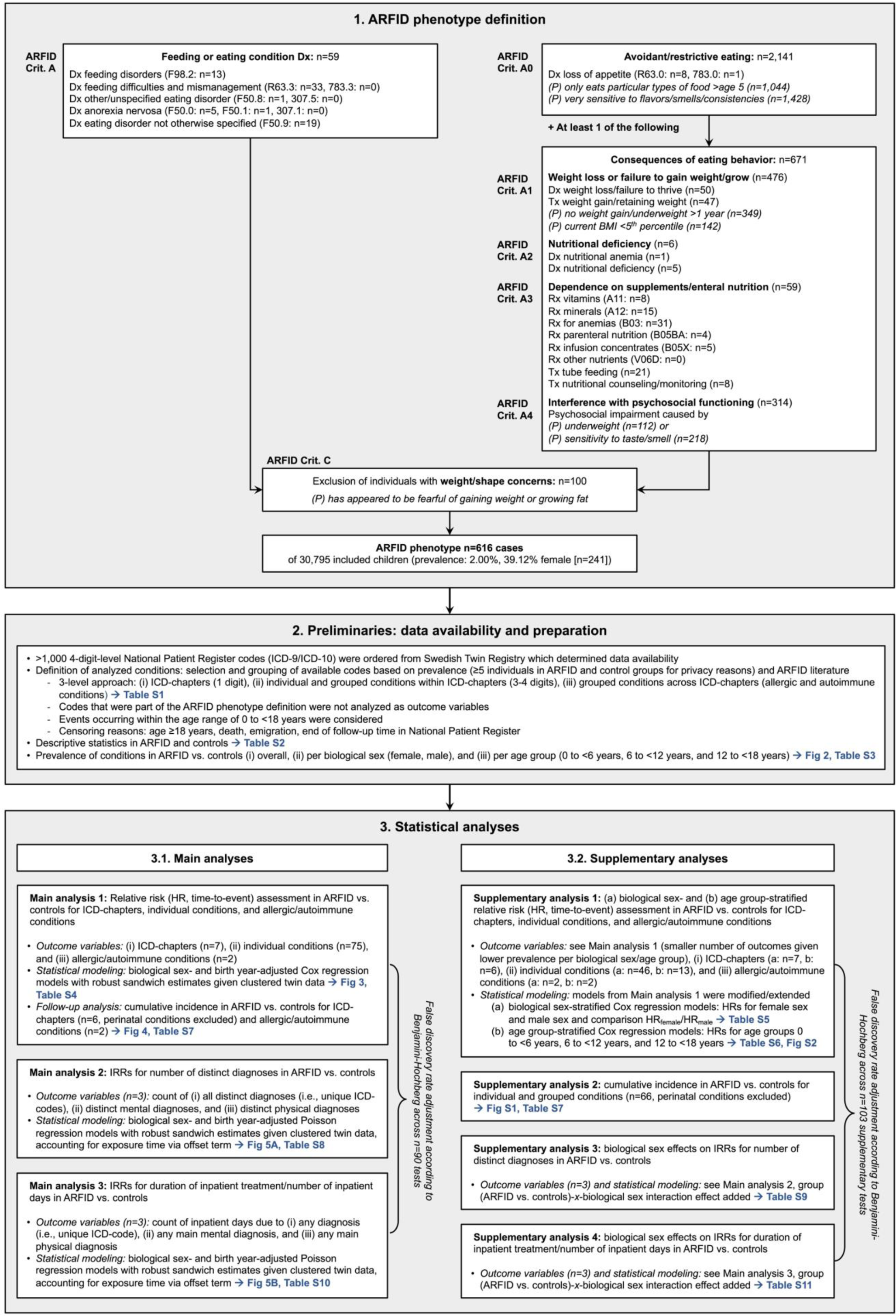
ARFID phenotype definition and statistical analysis plan. ARFID phenotype definition (1.) is based on Diagnostic and Statistical Manual of Mental Disorders 5^th^ Edition (DSM-5) criteria. Available diagnoses, procedures, and prescribed drugs between twin age 6-12 years were included (data sources: Child and Adolescent Twin Study in Sweden [CATSS], National Patient Register [NPR], Prescribed Drug Register [PDR]). Elements in *italics*, designated as *(P)*, were parent-reported (CATSS at twin age 9 or 12 years). Numbers (n) of individuals who met the specified diagnostic criteria/elements at each step of the ARFID phenotype definition are stated, including overall prevalence of the ARFID phenotype out of all individuals in the study and proportion of girls among individuals with the ARFID phenotype. *Example for ARFID phenotype identification: A child was identified with ARFID if (1) parents reported that their 9-year-old child only had only eaten particular types of food after the age of 5 (criterion A: avoidant/restrictive eating), (2) the child had been diagnosed with nutritional deficiency between age 6 and 12 years (criterion A: clinical consequence of avoidant/restrictive eating), and (3) parents reported that their 9-year-old child had not appeared fearful of gaining weight (criterion C).* Analyzed conditions were ICD-chapters (1-digit codes), individual and grouped conditions within ICD-chapters (3-/4-digit codes) as well as grouped conditions across ICD-chapters (allergic and autoimmune conditions). Statistical analyses (3.) were structured in main analyses (relative and absolute risk assessments for analyzed conditions in ARFID vs. controls, Poisson regressions analyzing the number of distinct diagnoses [unique ICD-codes], and days of inpatient treatment per individual twin in ARFID vs. controls) and supplementary analyses (sex-specific and age group-specific assessments). Multiple testing adjustment via the false discovery rate (Benjamini-Hochberg^21^) was applied across all main analyses and separately across all supplementary analyses. Please note that cumulative incidence analyses (under 3.1. and 3.2.) were descriptive and did not contribute to the overall number of tests/false discovery rate adjustment. Age group-stratified analyses (under 3.2., supplementary analysis 1B) produced 2 tests (age group comparisons) per outcome variable. *Abbreviations: ARFID, avoidant restrictive food intake disorder; Dx, diagnosis; HR, hazard ratio, ICD-9/10, International Statistical Classification of Diseases and Related Health Problems Ninth/Tenth Revision; IRR, incidence rate ratio; Rx, prescription; Tx, treatment/procedure*.

### Outcomes

NPR diagnostic codes (>1,000 individual 3-/4-digit ICD-9/-10-codes) for the time range 1992–2016 were used to define the mental and somatic conditions that were analyzed as study outcomes (ARFID phenotype-defining codes were not analyzed as outcomes; **Figure 1**). Relevant conditions were defined via selection and grouping of the available ICD-codes according to clinical context and symptom overlap, previous ARFID literature, and prevalence in our sample. For a condition to be analyzed as an outcome, we required at least n=5 individuals with the condition in both the ARFID and the control group to maintain privacy. Outcome definition followed a 3-level approach: (i) ICD-chapters (1-digit-code level, e.g., endocrine/metabolic disorders [chapter E]; 7 outcomes), (ii) individual and grouped conditions within ICD-chapters (3-/4-digit-code level, e.g., diabetes mellitus and thyroid conditions [within chapter E]; 75 outcomes), and (iii) grouped allergic and autoimmune conditions across ICD-chapters (2 outcomes; see **Table S2** for detailed descriptions and ICD-code allocations for all outcomes).

Further study outcomes included number of *all* distinct diagnoses (i.e., unique ICD-codes) per person (as well as number of distinct *mental* and *somatic* diagnoses separately), and number of inpatient days per person due to any diagnosis (as well as due to any *mental* and any *somatic* diagnosis separately, 6 outcomes; **Figure 1**). Only conditions occurring between birth and 18^th^ birthday, prior to potential death, emigration, or end of follow-up (2016) qualified as outcomes (age at end of follow-up: *M*=15.35, *SD*=3.15).

### Statistical analyses

Statistical analyses were performed in R version 4.2.2.^20^ All tests were 2-tailed; statistical significance was indicated by the Benjamini-Hochberg false discovery rate (FDR)^21^-adjusted threshold *α*_FDR_=0.0343 (correction across all 90 main outcomes). Correlations among twin pairs were statistically addressed by using cluster-robust standard errors and confidence intervals (CIs). All models accounted for exposure/follow-up time and potential censoring during this time.

**Figure 1** provides an overview of all conducted analyses. Lifetime prevalence of outcomes in ARFID and controls was computed as *n_diagnosed_/n_total_* overall, per female/male sex, and per age group (birth to <6 years, 6 to <12 years, and 12 to <18 years). For main analysis 1, we performed time-to-event Cox proportional hazards regression models to assess relative risks of the 84 analyzed conditions in ARFID vs. controls via hazard ratios (HRs) and their cluster-robust 95% CIs. The following specifications applied:

- Twin age was the underlying time scale (birth to 18^th^ birthday),
- First diagnosis of the analyzed condition constituted the event,
- ARFID phenotype served as time-invariant predictor (i.e., presence of the ARFID phenotype was modeled as a consistent exposure from birth to 18^th^ birthday),
- Adjustment for sex assigned at birth (female/male) and birth year (range: 1992–2008) as categorical covariates,
- Supplementary sex- and age group-stratified Cox regression models were implemented to assess and compare sex- and age group-specific HRs in ARFID vs. controls (e.g., HRs in girls with ARFID vs. female controls; age groups: early childhood, middle/late childhood, adolescence).

As a descriptive follow-up analysis, we calculated absolute risk via cumulative incidence of chapter-level (except for perinatal) and grouped allergic/autoimmune conditions in ARFID vs. controls from birth to 18^th^ birthday (supplementary analysis for individual conditions). Kaplan-Meier estimation was used after matching n=10 unexposed children to each child with ARFID based on sex and birth year. Thus, we obtained the proportion of children per group (ARFID/controls) diagnosed with any analyzed condition prior to a specific age while accounting for censoring and covariates.

To estimate treatment needs in ARFID, we computed incidence rate ratios (IRRs) and their cluster-robust 95% CIs for the count of *all* distinct, all distinct *mental*, and all distinct *somatic* ICD-diagnoses per person in ARFID vs. controls (main analysis 2). Likewise, we computed IRRs for the count of inpatient days due to *any*, any primary *mental*, and any primary *somatic* ICD-diagnosis in ARFID vs. controls (main analysis 3). We employed Poisson regression models, adjusted for sex and categorical birth year and including an offset term for exposure time.

Supplementary analyses examined potential interaction effects of group (ARFID/controls) and sex on number of distinct diagnoses and inpatient days.

## Results

### Sample characteristics and prevalence of outcomes

Of children and adolescents meeting our ARFID phenotype criteria (n=616, 2.00% of the total sample), 60.88% (n=375) were male (**Table S1**). Within the ARFID group, 87.18% of children had one or more registered diagnoses (excluding any diagnoses that were part of the ARFID phenotype), compared with 73.03% in children without ARFID. In children with ARFID, the prevalence of any diagnosis within an ICD-chapter was (in descending order): J: Respiratory conditions (53.08%); F: Mental conditions (35.88%); P: Perinatal conditions (34.42%); K: Digestive conditions (34.09%); Q: Congenital conditions (22.40%); E: Endocrine/metabolic conditions (17.69%); and I: Circulatory conditions (4.06%; **Figure 2, Table S3**). This order of ICD-chapter prevalence was similar in controls, except for F: Mental conditions, which were second most common in children with ARFID but only the fifth most common conditions in controls. Within F: Mental conditions, the most common conditions in children with ARFID were ADHD (17.5%), autism (13.8%), and intellectual disability (9.1%). The most common *somatic* conditions in children with ARFID were: acute upper respiratory infections (30.36%), fetal growth retardation (28.73%), allergic conditions (27.06%), asthma (20.78%), acute lower respiratory infections (18.18%), chronic upper respiratory disorders (15.26%), neonatal jaundice (14.77%), and constipation (14.12%).

**Figure 2.**
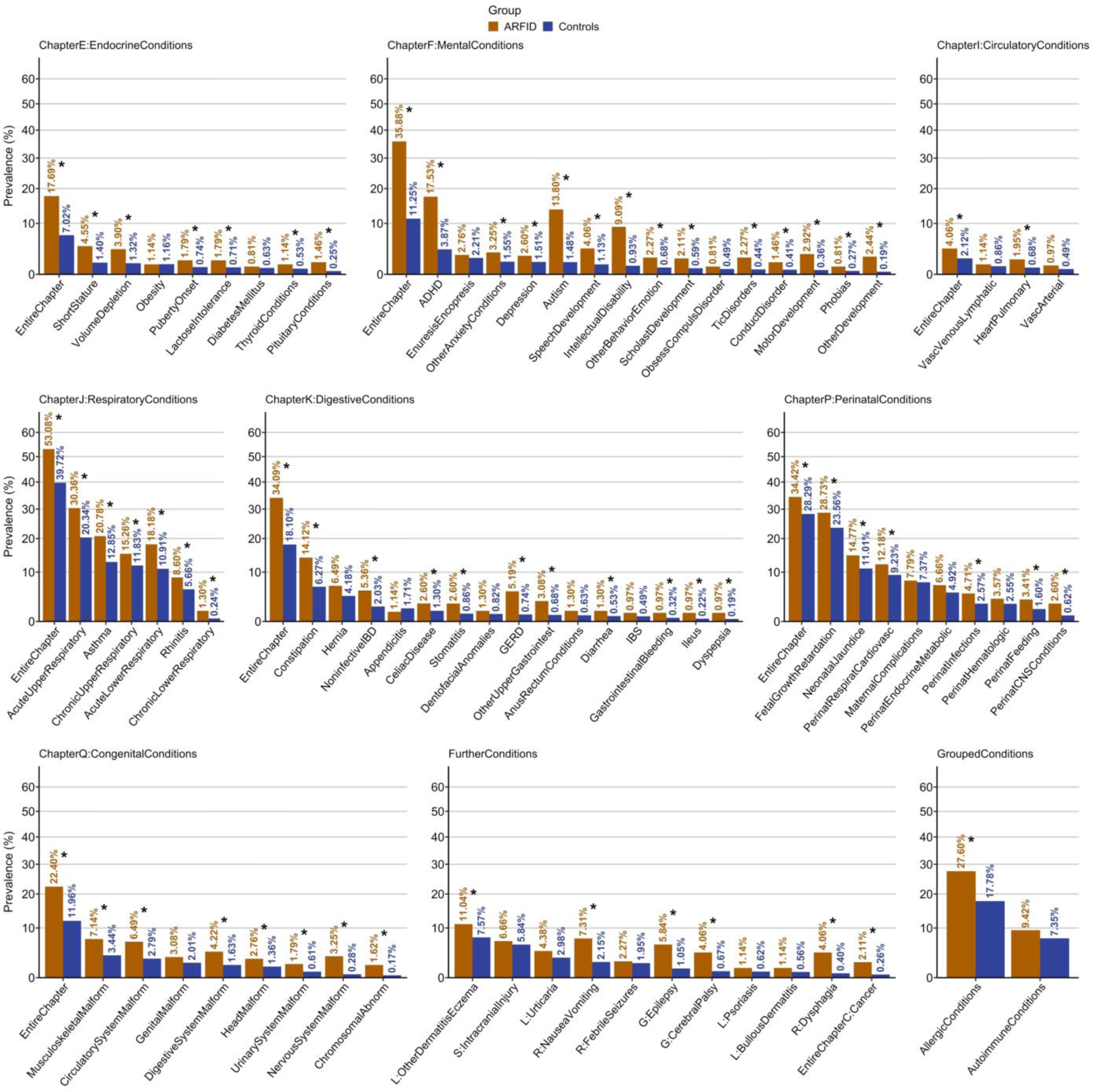
Prevalence of co-existing conditions in children and adolescents with ARFID vs. controls. Bar plots for prevalence of each analyzed condition (n_diagnosed_/n_total_, %) in ARFID and controls (y-axis logarithmically scaled). Conditions are sorted in descending order of prevalence in controls within ICD-chapters, the group of further conditions, and grouped allergic and autoimmune conditions. Conditions for which hazard ratios were significantly different (i.e., increased) in ARFID compared with controls according to the base Cox regression model at the false discovery rate-adjusted threshold *α*_FDR_=0.0343 are marked by an asterisk (*). Please note that some labels were collapsed to indicate conditions in a space-efficient manner (please consult Table S2 for details). *Abbreviations: ADHD, attention deficit hyperactivity disorder; ARFID, avoidant restrictive food intake disorder; CNS, central nervous system; GERD, gastro-esophageal reflux disorder; IBD, inflammatory bowel disease; IBS, irritable bowel syndrome*.

### Relative and absolute risks

As anticipated, relative risks for the majority of analyzed conditions were significantly increased in children with ARFID compared with controls (**Figure 3, Table S4**). Cox regressions yielded HRs>1 in ARFID vs. controls for all seven included ICD-chapters, with highest relative risk of mental conditions (HR=3.90 [CI_95%_=3.36-4.52]), followed by endocrine/metabolic (HR=2.73 [CI_95%_=2.21-3.36]) and digestive (HR=2.03 [CI_95%_=1.76-2.35]) conditions. Children with ARFID had significantly elevated risk of 55 of 75 conditions (73.33%) *within* ICD-chapters, with highest HRs for mixed specific/other/unspecified developmental disorders (F83/F88/F89, e.g., developmental agnosia), nervous system malformations; dysphagia, intellectual disability, autism, cancer, motor development impairment, gastroesophageal reflux disease (GERD), cerebral palsy, epilepsy, upper gastrointestinal conditions, ADHD, and tic disorders. In contrast, ARFID was *not* associated with significantly increased risk of 20 of 75 conditions *within* ICD-chapters, including irritable bowel syndrome (IBS), obsessive-compulsive disorder (OCD), diabetes, and obesity (**Figure 3**). While the risk of allergic conditions was larger in ARFID than controls, the risk of autoimmune conditions was not significantly increased (**Figure 3**).

**Figure 3.**
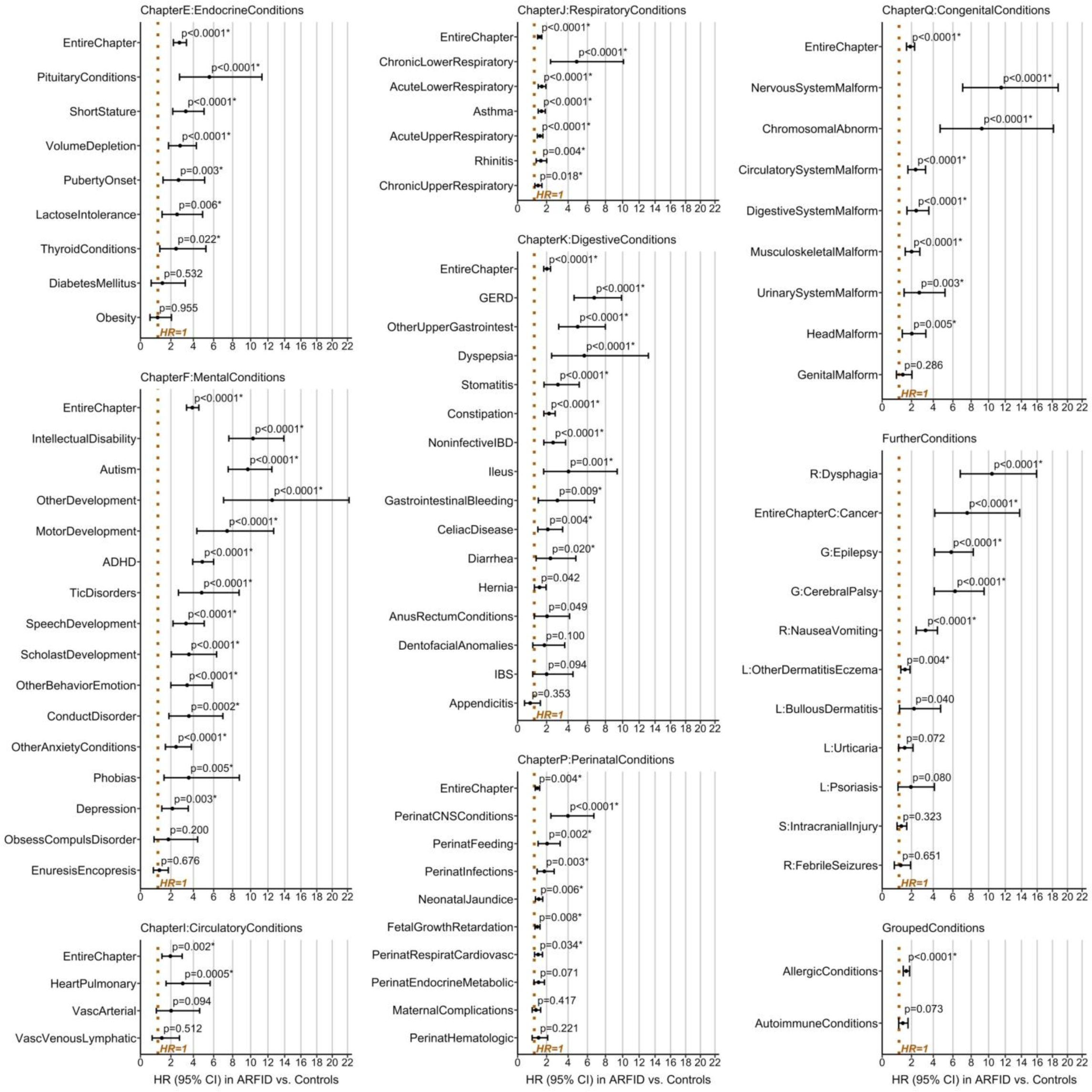
Hazard ratio plots for co-existing conditions in children and adolescents with ARFID vs. controls. Relative risk via hazard ratio (HR) in ARFID vs. controls, its cluster-robust 95% confidence interval (95% CI), and p-value are displayed for each analyzed condition (x-axis logarithmically scaled). Conditions for which risks were significantly different (i.e., increased) in ARFID compared with controls at the false discovery rate-adjusted threshold *α*_FDR_=0.0343 are marked by an asterisk (*). Estimates and statistics were obtained with the base cox regression model (predictor: group [ARFID vs. controls]; covariates: sex [female/male], birth year [1992–2008, factorized]; robust sandwich estimates given clustered twin data). Conditions are sorted in descending order of the lower limit of the 95% CI of HRs within ICD-chapters, the group of further conditions, and grouped allergic and autoimmune conditions. Please note that some labels were collapsed to indicate conditions in a space-efficient manner (please consult Table S2 for details). *Abbreviations: ADHD, attention deficit hyperactivity disorder; ARFID, avoidant restrictive food intake disorder; CNS, central nervous system; GERD, gastro-esophageal reflux disorder; IBD, inflammatory bowel disease; IBS, irritable bowel syndrome*.

Inverted Kaplan-Meier curves in **Figure 4** illustrate steeper increase in cumulative incidence with increasing age in ARFID vs. controls for all ICD-chapter-level conditions; primarily for mental conditions (steepest increase in ARFID relative to controls between age 7 and 11 years), respiratory conditions (steepest increase from birth to age 4 years with subsequent flattening), and endocrine, digestive, and allergic conditions (more steadily diverging curves in ARFID vs. controls with increasing age, **Figure 4**). In addition, cumulative incidence diverged substantially in ARFID vs. controls for several individual conditions, including ADHD, autism, intellectual disability, asthma, GERD, and epilepsy (**Figure S1**). However, for grouped autoimmune conditions (**Figure 4**) and other individual conditions such as diabetes, OCD, IBS, and febrile seizures (**Figure S1**, **Table S5**), Kaplan-Meier curves were overlapping indicating comparable absolute risks with increasing age in children with ARFID and controls.

**Figure 4.**
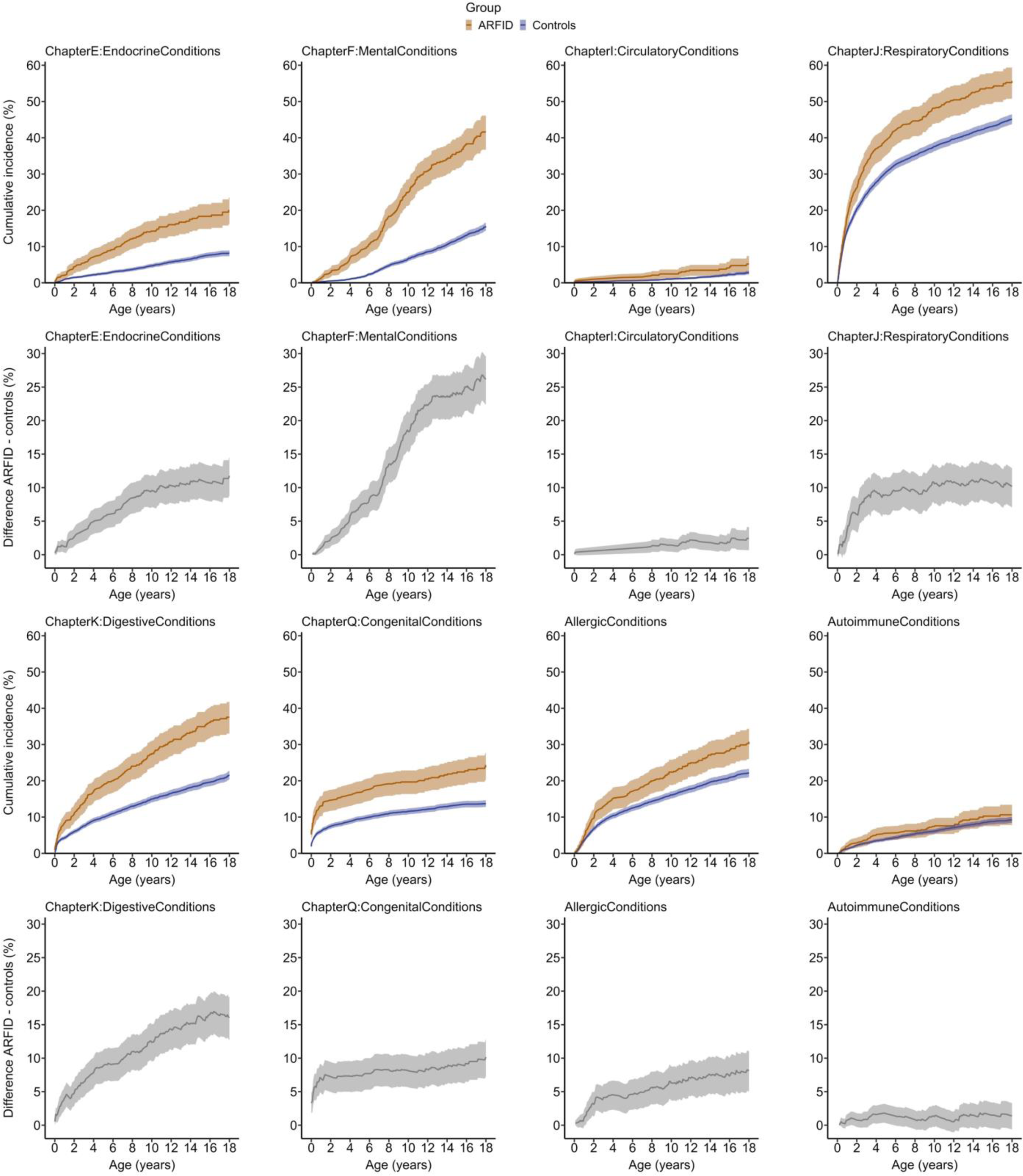
Cumulative incidence plots for co-existing conditions in children and adolescents with ARFID vs. controls. Absolute risk via cumulative incidence (%) of having a specific condition (ICD-chapters and grouped allergic and autoimmune conditions) and lower and upper limits of its cluster-robust 95% confidence interval are plotted over age (0 to <18 years) in ARFID vs. controls. As a follow-up analysis, difference (%) in cumulative incidence was computed as ARFID - controls and plotted for each condition below the corresponding cumulative incidence plot. Perinatal conditions were excluded given their occurrence during the perinatal period and lack of age-related dynamics. Cumulative incidence was estimated using the Kaplan-Meier method (1 - Kaplan-Meier survival estimate, accounting for censoring) after creating a modified analysis sample by matching n=10 unexposed individuals to each (n=1) individual with ARFID, stratified by sex and birth year. *Abbreviation: ARFID, avoidant restrictive food intake disorder*.

### Number of distinct diagnoses and inpatient treatment duration

Children with ARFID presented with a significantly higher number of distinct diagnoses per person (M=6.27 [SE=0.28]) compared with controls (M=2.78 [SE=0.02]; **Figure 5, Table S6**). In particular, incidence rate of number of distinct *mental* diagnoses was almost fivefold higher in ARFID than controls (IRR=4.65 [CI_95%_=3.99-5.42]), whereas incidence rate of distinct *somatic* diagnoses was twice as high in ARFID (IRR=1.98 [CI_95%_=1.79-2.19]).

**Figure 5.**
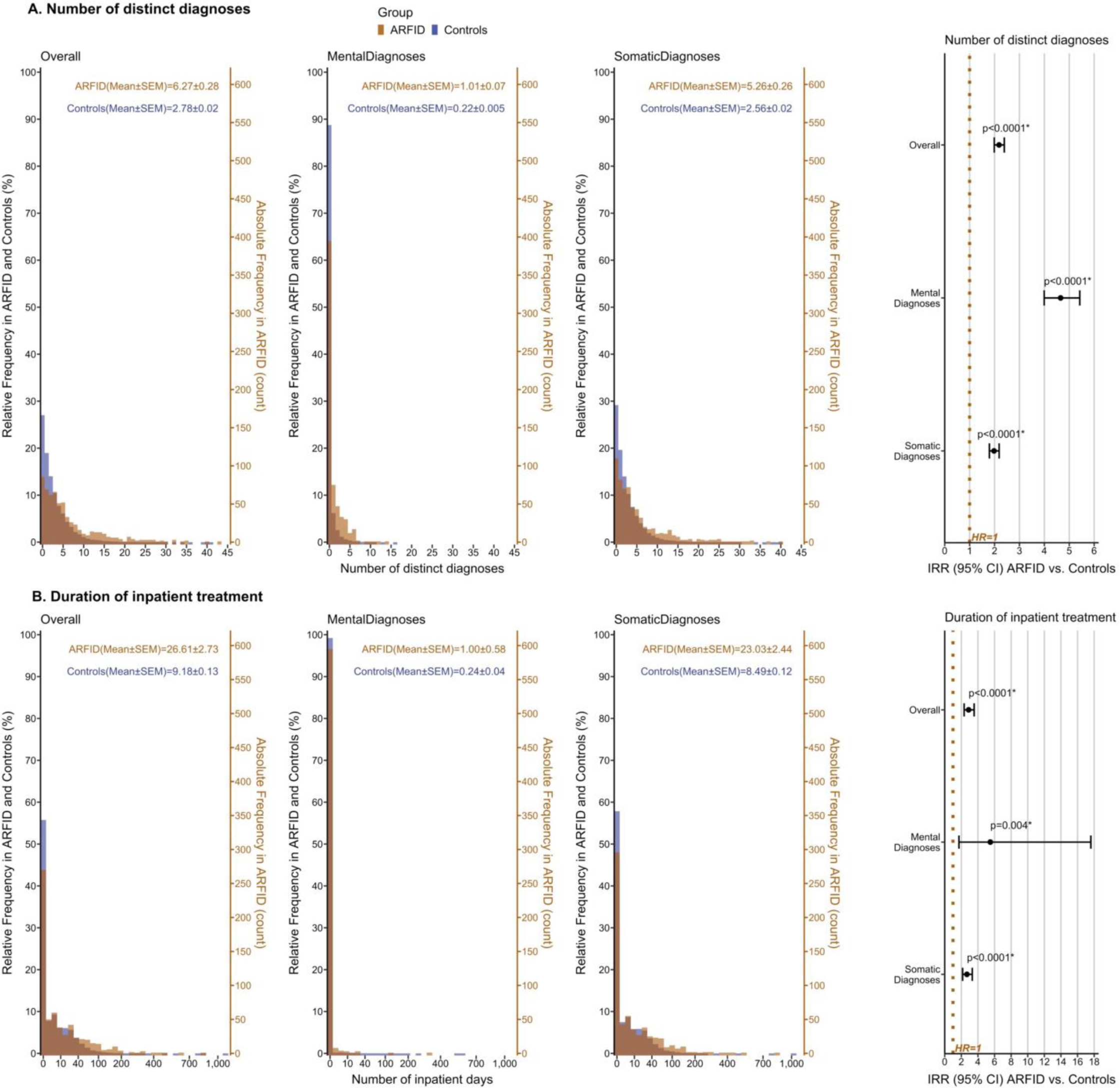
A. Number of all distinct, distinct mental, and distinct somatic ICD-diagnoses in ARFID vs. controls. B. Duration of hospitalization/inpatient treatment due to any, any mental, and any somatic ICD-diagnosis in ARFID vs. controls. Histograms display absolute frequency (count) and relative frequency (%) of A. the number of distinct diagnoses (i.e., unique ICD-codes) and B. the number of inpatient days per individual twin in ARFID vs. controls (x-axis in B. logarithmically scaled). Mean and standard error of the mean (SEM) are provided as descriptive statistics. Poisson regression models (predictor: group [ARFID vs. controls]; covariates: sex [female/male], birth year [1992–2008, factorized]; robust sandwich estimates given clustered twin data; offset term to account for exposure time) were applied to estimate incidence rate ratios (IRRs) in ARFID vs. controls in A. and B., their cluster-robust 95% confidence intervals (95% CI), and p-values (x-axis logarithmically scaled). Significantly different (i.e., increased) incidence rates in ARFID compared with controls at the false discovery rate-adjusted threshold *α*_FDR_=0.0343 are marked by an asterisk (*). *Abbreviation: ARFID, avoidant restrictive food intake disorder*.

Regarding inpatient treatment duration, children with ARFID were hospitalized for substantially more days per person (M=26.61 [SE=2.73]) than controls (M=9.18 [SE=0.13]; IRR=2.88 [CI_95%_=2.34-3.55]). Although, overall, the majority of inpatient days was due to *somatic* diagnoses, risk increase of inpatient days in the ARFID group was most substantial for *mental* diagnoses leading to hospitalization (IRR=5.50 [CI_95%_=1.72-17.60]; **Figure 5, Table S7**).

### Roles of sex and age group

Sex-stratified Cox regressions yielded differences in HRs (ARFID vs. controls) that did not survive FDR-adjustment for the following conditions: short stature, motor development, congenital conditions (chapter Q), and chronic upper respiratory conditions, for which girls had higher relative risk than boys, whereas the risk was higher in boys than girls for anxiety disorders (**Table S8**). Furthermore, we did not detect any effects of sex on the number of distinct diagnoses (**Table S9**) or the number of inpatient days (**Table S10**).

ARFID was associated with an increased relative risk of being diagnosed with a mental condition (chapter F) at a younger age (6 to <12 years vs. 12 to <18 years; **Table S11**). The age-varying course of HRs for mental conditions is illustrated in **Figure S2**. Moreover, prior to FDR-adjustment, autism, ADHD, and endocrine/metabolic conditions (chapter E) tended to be diagnosed *earlier* in children with ARFID than in controls (HR 6 to <12 years > HR 12 to <18 years), whereas some dermatological conditions (dermatitis, eczema) tended to be diagnosed *later* in children with ARFID compared with controls (HR 6 to <12 years < HR 12 to <18 years; **Table S11**).

## Discussion

In this study of children and adolescents with and without the ARFID phenotype, we found an elevated risk of co-existing somatic and particularly mental conditions in individuals with ARFID. Relative risks of all ICD-chapter-level conditions, grouped allergic conditions, and the majority of individually analyzed conditions were statistically significantly increased in ARFID vs. controls. For instance, individuals with ARFID had a tenfold risk of autism, intellectual disability, and dysphagia; and a sixfold risk of cerebral palsy and epilepsy. Furthermore, treatment needs, indicated by number of distinct diagnoses and hospitalization duration, were substantially increased in ARFID vs. controls, with mental disorders primarily contributing to group differences. On average, we recorded ≈6 distinct diagnoses and ≈26 inpatient days per child with ARFID.

Overall, the highest relative risk increase was recorded for *mental* conditions: ≈36% of children with ARFID had a diagnosed mental condition (compared with ≈11% in controls). In particular, we observed a particularly strong risk increase in ARFID vs. controls for a variety of NDCs, including intellectual disability, autism, mixed specific/other/unspecified developmental disorders, ADHD, tic disorders, and motor development disorder. Previous studies focused mostly on autism and ADHD and, thus, did not show the same broad range of NDCs in individuals with ARFID that our study yielded. Nonetheless, the most common mental conditions in our sample were ADHD (17.5%), autism (13.8%), and intellectual disability (9.1%), which supports findings from smaller-scale clinical studies where these NDCs occurred in ARFID with even higher prevalence.^13,22^ The relatively lower NDC prevalence in ARFID here may be explained by our general population sample with overall less symptom severity.

Regarding *somatic* conditions, we found the highest risk increase in ARFID vs. controls for gastrointestinal disorders (specifically GERD, upper gastrointestinal disorders, and dysphagia); neurological disorders (cerebral palsy and epilepsy); cancer, and nervous system malformations. The co-occurrence of ARFID and gastrointestinal disorders has repeatedly been highlighted.^6,23^ Digestive symptoms, sensory aversion, fear of aversive gastrointestinal consequences, and food avoidance can reinforce and perpetuate each other, thereby contributing to diagnostic overshadowing.

Children with ARFID also had a higher risk of respiratory conditions. This in line with two recent studies based on small samples and case reports showing an elevated prevalence of asthma^11^ and other respiratory complications in ARFID (e.g., aspiration pneumonia, pneumothorax, and non-tuberculosis mycobacteria infections).^24^ We also observed increased risk of combined allergic conditions in ARFID, including respiratory, gastrointestinal, and dermatological allergic conditions, which could contribute to food sensitivities and/or fear of negative consequences of eating. The available level of detail of NPR diagnostic codes did not allow us to specify food allergies, which have been shown to be common in ARFID.^11^

Our finding of increased risk of cancer in ARFID (overall low prevalence: n=13 in ARFID, n=79 in controls) needs to be interpreted with caution, given limited statistical power and our epidemiological study design, which broadly mapped co-existing conditions in ARFID. Therefore, we could not clearly discern whether certain medical conditions should be considered cause, comorbidity, or consequence of ARFID. Some conditions described here may potentially explain avoidant/restrictive food intake and its consequences. For instance, we cannot exclude that for some children, adverse effects of cancer treatment (e.g., appetite loss and associated weight loss) may have mimicked ARFID symptoms.

We did *not* find a significantly increased risk of OCD in children ARFID, which may indicate that the association is weaker than we had hypothesized, although previous evidence is scarce. In a general population sample using a similar procedure to identify the ARFID phenotype (n=183 with ARFID), significantly more OCD symptoms were reported for the ARFID group.^25^ However, the effect size was small and the statistical power was greater due to the use of a parent-reported continuous measure (rather than clinical diagnoses in the current study).

In contrast to previous epidemiological evidence on strong bidirectional relationships of autoimmune conditions and eating disorders,^26^ the lack of association between the ARFID phenotype and grouped autoimmune conditions might evolve from demographic sample characteristics like the upper age boundary of follow-up in this study (<18 years; mean age of follow-up 15.3 years). For example, the median age at diagnosis of type I diabetes is 24 years^27^ and of inflammatory bowel disease 20–30 years.^28^ Our twins may not yet have passed through the age of risk of developing these autoimmune conditions. Importantly, due to a prevalence of n<5 in the ARFID group (**Table S2**), we could not examine associations between ARFID and autoimmune pediatric acute-onset neuropsychiatric syndrome following group A streptococcus infection (PANS/PANDAS), even though evidence of the interplay of PANS/PANDAS, OCD, and ARFID exists.^29^

In contrast to controls, children with ARFID were more likely to be diagnosed with a mental disorder in late childhood (6 to <12 years) than in early adolescence (12 to <18 years). In fact, cumulative incidence curves demonstrated the steepest increase in mental conditions in ARFID relative to controls between 7 and 11 years of age, which aligns with the peak age at first NDC diagnosis in the literature.^30^ Sex-specific relative risk differences between ARFID and controls did not survive multiple testing correction in this study. To our knowledge, this is the first study on links between sex and comorbidity in ARFID. Previously, ARFID profiles (fear of aversive consequences, lack of interest, and sensory-based avoidance) were found to be independent of sex.^22^ Another study found sex differences in NDC comorbidity with in children diagnosed with a feeding and eating disorder before age 3.^31^

This study has several strengths. We investigated ARFID-related mental and somatic comorbidities in the largest reported sample of children and adolescents with the ARFID phenotype to date. Previous evidence on comorbidities in ARFID is based on small-scale, frequently uncontrolled, clinical studies in subpopulations. Thus, our large-scale epidemiological data, including clinical diagnoses from high-quality sources and different clinical settings,^18,19^ enhances knowledge and generalizability of the complex clinical presentations of ARFID. Furthermore, this study covers a broad range of medical conditions in ARFID (n=84), several of which had not been previously examined (e.g., various NDCs including disorders of speech, motor, scholastic, and general development; fine-grained specification of respiratory, perinatal, and congenital conditions). Additionally, the indices of treatment needs (number of distinct per-person diagnoses, hospitalization duration) provide novel insight into the complex healthcare needs associated with ARFID.

The following limitations should be considered. Given this is an observational study with the aim to characterize the health care needs of people with ARFID, our findings highlight co-occurrence. ARFID was not treated as a time varying exposure and co-occurring conditions could have been diagnosed before or after the actual onset of ARFID symptoms, which was unknown to us. Additionally, we could not infer whether certain conditions might potentially have accounted for avoidant/restrictive eating or impairment of nutrition and weight development. Therefore, we cannot rule out that the more inclusive *pediatric feeding disorder* phenotype would have better matched some children identified with ARFID here.^32^ Future studies should assess the impact of treating ARFID as time varying or not (using larger sample sizes). Second, due to its definition, our ARFID phenotype reflects mainly cases that include a sensory-based avoidance component, which typically is associated with selective eating^33^ and might have biased our results towards higher co-occurrence of NDCs. Third, we relied on NPR as the source of diagnostic information; however, NPR does not contain diagnoses given in primary health care. Other conditions might not have come to medical attention (no NPR diagnosis). Lastly, despite a sample size of >30,000, some analyses for conditions with low prevalence in our sample (e.g., OCD) as well as sex-/age group-specific analyses remained underpowered. Even larger data sets and longer follow-up periods will be needed to obtain well-powered analyses.

Here, we provide the broadest and most detailed systematic evidence of co-existing mental and somatic conditions in the largest sample of children with ARFID and controls to date. We demonstrate that the ARFID phenotype is associated with higher prevalence and increased risk of multiple comorbidities such as a variety of neurodevelopmental, gastrointestinal, endocrine/metabolic, and neurological disorders. Children with ARFID presented with greater treatment needs than controls. Our findings suggest heterogenous and complex clinical presentations of ARFID in childhood and adolescence, which underscore the critical importance of raising awareness and competence in providers from multiple specialties (e.g., internal medicine, endocrinology, gastroenterology, oncology, general pediatrics, general practice, psychiatry) to improve detection, diagnosis, and treatment of ARFID.

## Supplement 1: Supplementary Material

Supplementary Material associated with this article can be found in the online version at [URL].

## Supplement 2: Analysis R script

Full de-identified analysis R script available on Github (open access): https://github.com/MarieLouisW/Co-existing-mental-and-somatic-conditions-in-avoidant-restrictive-food-intake-disorder.git

## Disclosures

### Authors’ contributions

Conceptualization: Marie-Louis Wronski, Lisa Dinkler, Nadia Micali, Cynthia M. Bulik

Data curation: Marie-Louis Wronski, Lisa Dinkler

Formal analysis: Marie-Louis Wronski, Lisa Dinkler

Funding acquisition: Lisa Dinkler

Investigation: Marie-Louis Wronski, Lisa Dinkler, Paul Lichtenstein, Sebastian Lundström, Henrik Larsson

Methodology: Marie-Louis Wronski, Lisa Dinkler, Ralf Kuja-Halkola, Mark J. Taylor

Project administration: Lisa Dinkler

Resources: Paul Lichtenstein, Sebastian Lundström, Henrik Larsson

Software: Marie-Louis Wronski, Ralf Kuja-Halkola, Miriam I. Martini

Supervision: Lisa Dinkler

Validation: Lisa Dinkler, Elin Hedlund, Nadia Micali

Visualization: Marie-Louis Wronski, Lisa Dinkler

Writing – original draft: Marie-Louis Wronski, Lisa Dinkler

Writing – review & editing: All authors

### Data sharing

Swedish register data are not publicly available and cannot be shared. However, for transparency of the applied analyses, the authors share their analysis script (available on Github [open access]: https://github.com/MarieLouisW/Co-existing-mental-and-somatic-conditions-in-avoidant-restrictive-food-intake-disorder.git).

## Acknowledgements

We gratefully acknowledge the contribution of the participants in the Child and Adolescent Twin Study in Sweden (CATSS) and their families. We thank the Swedish Twin Registry for granting us data access. The Swedish Twin Registry is managed by Karolinska Institutet and receives funding through the Swedish Research Council under grant 2017-00641.

## References

1. Dinkler L, Wronski ML, Lichtenstein P, Lundstrom S, Larsson H, Micali N, et al. Etiology of the Broad Avoidant Restrictive Food Intake Disorder Phenotype in Swedish Twins Aged 6 to 12 Years. JAMA Psychiatry. 2023;80(3):260–9.

2. Katzman DK, Spettigue W, Agostino H, Couturier J, Dominic A, Findlay SM, et al. Incidence and Age- and Sex-Specific Differences in the Clinical Presentation of Children and Adolescents With Avoidant Restrictive Food Intake Disorder. JAMA Pediatr. 2021;175(12):e213861.

3. American Psychiatric Association. Diagnostic and Statistical Manual of Mental Disorders, Fifth Edition. Arlington, VA: American Psychiatric Association; 2013.

4. Harshman SG, Jo J, Kuhnle M, Hauser K, Murray HB, Becker KR, et al. A Moving Target: How We Define Avoidant/Restrictive Food Intake Disorder Can Double Its Prevalence. J Clin Psychiatry. 2021;82(5).

5. Nicholas JK, van Tilburg MAL, Pilato I, Erwin S, Rivera-Cancel AM, Ives L, et al. The diagnosis of avoidant restrictive food intake disorder in the presence of gastrointestinal disorders: Opportunities to define shared mechanisms of symptom expression. Int J Eat Disord. 2021;54(6):995–1008.

6. Weeks I, Abber SR, Thomas JJ, Calabrese S, Kuo B, Staller K, et al. The Intersection of Disorders of Gut-Brain Interaction With Avoidant/Restrictive Food Intake Disorder. J Clin Gastroenterol. 2023.

7. Cifra N, Lomas JM. Differentiating Eosinophilic Esophagitis and Eating/Feeding Disorders. Pediatrics. 2022;149(4).

8. Rajendram R, Psihogios M, Toulany A. Delayed diagnosis of avoidant/restrictive food intake disorder and autism spectrum disorder in a 14-year-old boy. Clin Case Rep. 2021;9(6):e04302.

9. Sanchez-Cerezo J, Nagularaj L, Gledhill J, Nicholls D. What do we know about the epidemiology of avoidant/restrictive food intake disorder in children and adolescents? A systematic review of the literature. Eur Eat Disord Rev. 2023;31(2):226–46.

10. Kambanis PE, Kuhnle MC, Wons OB, Jo JH, Keshishian AC, Hauser K, et al. Prevalence and correlates of psychiatric comorbidities in children and adolescents with full and subthreshold avoidant/restrictive food intake disorder. Int J Eat Disord. 2020;53(2):256–65.

11. Aulinas A, Marengi DA, Galbiati F, Asanza E, Slattery M, Mancuso CJ, et al. Medical comorbidities and endocrine dysfunction in low-weight females with avoidant/restrictive food intake disorder compared to anorexia nervosa and healthy controls. Int J Eat Disord. 2020;53(4):631–6.

12. Becker KR, Mancuso C, Dreier MJ, Asanza E, Breithaupt L, Slattery M, et al. Ghrelin and PYY in low-weight females with avoidant/restrictive food intake disorder compared to anorexia nervosa and healthy controls. Psychoneuroendocrinology. 2021;129:105243.

13. Dinkler L, Yasumitsu-Lovell K, Eitoku M, Fujieda M, Suganuma N, Hatakenaka Y, et al. Early neurodevelopmental problems and risk for avoidant/restrictive food intake disorder (ARFID) in 4-7-year-old children: A Japanese birth cohort study. JCPP Advances. 2022;2(3):e12094.

14. Hambleton A, Pepin G, Le A, Maloney D, Touyz S, Maguire S. Psychiatric and medical comorbidities of eating disorders: findings from a rapid review of the literature. J Eat Disord. 2022;10(1):132.

15. Brosig L, Düplois D, Hiemisch A, Kiess W, Hilbert A, Schlensog-Schuster F, et al. Birth-related, medical, and diagnostic characteristics in younger versus older children with avoidant/restrictive food intake disorder (ARFID). J Eat Disord. 2023;11(1):190.

16. Anckarsater H, Lundstrom S, Kollberg L, Kerekes N, Palm C, Carlstrom E, et al. The Child and Adolescent Twin Study in Sweden (CATSS). Twin Res Hum Genet. 2011;14(6):495–508.

17. World Health Organization. International Statistical Classification of Diseases and Related Health Problems, Tenth Revision. Geneva: World Health Organization; 1992.

18. Ludvigsson JF, Andersson E, Ekbom A, Feychting M, Kim JL, Reuterwall C, et al. External review and validation of the Swedish national inpatient register. BMC Public Health. 2011;11:450.

19. Wettermark B, Hammar N, Fored CM, Leimanis A, Otterblad Olausson P, Bergman U, et al. The new Swedish Prescribed Drug Register--opportunities for pharmacoepidemiological research and experience from the first six months. Pharmacoepidemiol Drug Saf. 2007;16(7):726–35.

20. R Core Team. R: A language and environment for statistical computing. R Foundation for Statistical Computing, Vienna, Austria. https://www.R-project.org/. 2022.

21. Benjamini Y, Hochberg Y. Controlling the False Discovery Rate: A Practical and Powerful Approach to Multiple Testing. Journal of the Royal Statistical Society: Series B (Methodological). 1995;57(1):289–300.

22. Watts R, Archibald T, Hembry P, Howard M, Kelly C, Loomes R, et al. The clinical presentation of avoidant restrictive food intake disorder in children and adolescents is largely independent of sex, autism spectrum disorder and anxiety traits. EClinicalMedicine. 2023;63:102190.

23. Gibson D, Watters A, Mehler PS. The intersect of gastrointestinal symptoms and malnutrition associated with anorexia nervosa and avoidant/restrictive food intake disorder: Functional or pathophysiologic?-A systematic review. Int J Eat Disord. 2021;54(6):1019–54.

24. Nitsch A, Kearns M, Mehler P. Pulmonary complications of eating disorders: a literature review. J Eat Disord. 2023;11(1):12.

25. Sader M, Harris HA, Waiter GD, Jackson MC, Voortman T, Jansen PW, et al. Prevalence and Characterization of Avoidant Restrictive Food Intake Disorder in a Pediatric Population. JAACAP Open. 2023;1(2):116–27.

26. Hedman A, Breithaupt L, Hubel C, Thornton LM, Tillander A, Norring C, et al. Bidirectional relationship between eating disorders and autoimmune diseases. J Child Psychol Psychiatry. 2019;60(7):803–12.

27. Harris E. Large Number of People Diagnosed With Type 1 Diabetes After Age 30. JAMA. 2023;330(16):1516-.

28. Duricova D, Burisch J, Jess T, Gower-Rousseau C, Lakatos PL, ECCO-EpiCom OBo. Age-related differences in presentation and course of inflammatory bowel disease: an update on the population-based literature✩. Journal of Crohn’s and Colitis. 2014;8(11):1351–61.

29. Toufexis MD, Hommer R, Gerardi DM, Grant P, Rothschild L, D’Souza P, et al. Disordered eating and food restrictions in children with PANDAS/PANS. J Child Adolesc Psychopharmacol. 2015;25(1):48–56.

30. Plana-Ripoll O, Momen NC, McGrath JJ, Wimberley T, Brikell I, Schendel D, et al. Temporal changes in sex- and age-specific incidence profiles of mental disorders—A nationwide study from 1970 to 2016. Acta Psychiatr Scand. 2022;145(6):604–14.

31. Shan H, Li F, Zhang J, Wang H, Li J. Feeding and Eating Disorder and Risk of Subsequent Neurodevelopmental Disorders: A Population-Based Cohort Study. Front Pediatr. 2021;9:671631.

32. Noel RJ. Avoidant restrictive food intake disorder and pediatric feeding disorder: the pediatric gastroenterology perspective. Curr Opin Pediatr. 2023;35(5):566–73.

33. Bourne L, Mandy W, Bryant-Waugh R. Avoidant/restrictive food intake disorder and severe food selectivity in children and young people with autism: A scoping review. Dev Med Child Neurol. 2022;64(6):691–700.

