## Supplement 1: Supplementary Material for "Co-existing mental and somatic conditions in Swedish children with the avoidant restrictive food intake disorder phenotype"

### Table of contents


| Supplementary element | Content | Page |
|---|---|---|
| Table S1 | Descriptive statistics of the sample | 3 |
| Table S2 | Diagnostic classification of examined mental and somatic conditions in ARFID and controls | 4 |
| Table S3 | Prevalence of mental and somatic conditions in ARFID and controls overall, per sex, and per age group | 11 |
| Table S4 | Base Cox regression model estimates in ARFID vs. controls | 14 |
| Table S5 | Cumulative incidences of mental and somatic conditions before ages 6, 12, and 18 years in ARFID and controls | 16 |
| Table S6 | Number of distinct overall, all mental, and all somatic diagnoses in ARFID vs. controls | 19 |
| Table S7 | Number of inpatient days due to any, any mental, and any somatic diagnosis in ARFID vs. controls | 20 |
| Table S8 | Sex-stratified Cox regression model estimates in ARFID vs. controls | 21 |
| Table S9 | Sex effects on number of distinct overall, mental, and somatic diagnoses in ARFID vs. controls | 23 |
| Table S10 | Sex effects on number of inpatient days due to any, any mental, and any somatic diagnosis in ARFID vs. controls | 24 |
| Table S11 | Age group-stratified Cox regression model estimates in ARFID vs. controls | 25 |
| Figure S1 | Cumulative incidence plots in ARFID vs. controls for individual and grouped conditions | 26 |
| Figure S2 | Age-varying hazard ratios in ARFID vs. controls for ICD-chapter F: mental conditions | 28 |


### Definition of terms used in the Manuscript and Supplementary Material

ARFID, children and adolescents *with* the avoidant restrictive food intake disorder phenotype; controls, children and adolescents *without* the avoidant restrictive food intake disorder phenotype; sex, sex assigned at birth.

### Short labels for outcomes

Collapsed labels to indicate analyzed conditions in a space-efficient manner (applicable to all figures and tables in the Manuscript and Supplementary Material):

ChromosomalAbnorm, chromosomal abnormalities; Malform, malformation; NoninfectiveIBD, noninfective inflammatory bowel disease; ObsessCompulsDisorder, obsessive-compulsive disorder; OtherUpperGastrointest, other upper gastrointestinal conditions; Perinat, perinatal; PerinatCNSConditions, perinatal conditions of the central nervous system; PerinatRespiratCardiovasc, perinatal respiratory and cardiovascular conditions; ScholastDevelopment, scholastic development; VascArterial, vascular disorders of the arterial system; VascVenous, vascular disorders of the venous system.

**Table S1. Descriptive statistics of the sample**

| **Characteristic** n (%) | **1. Overall** | **2. ARFID** | **Controls** | **3. ARFID** | | **4. Controls** | | **Test statistics** | | |
|---|---|---|---|---|---|---|---|---|---|---|
| | | **Overall** | **Overall** | **Female** | **Male** | **Female** | **Male** | **Test** | $\chi^2$ | **p-value** |
| *A. Full sample* | 30,795 (100.00%) | 616 (2.00%) | 30,179 (98.00%) | 241 (39.12%) | 375 (60.88%) | 14,922 (49.44%) | 15,257 (50.56%) | Difference in sex distribution in ARFID vs. controls (overall) | 25.32 | <0.0001 |
| *B. Age at CATSS assessment* | | | | | | | | | | |
| - 9 years | 24,314 (100.00%) | 512 (2.11%) | 23,802 (97.89%) | 203 (39.65%) | 309 (60.35%) | 11,816 (49.64%) | 11,986 (50.36%) | Difference in CATSS assessment age in ARFID vs. controls (overall) | 6.30 | 0.012 |
| - 12 years | 6,481 (100.00%) | 104 (1.60%) | 6,377 (98.40%) | 38 (36.54%) | 66 (63.46%) | 3,106 (48.71%) | 3,271 (51.29%) | | | |
| *C. Birth year* | | | | | | | | | | |
| - 1992-1996 | 9,469 (100.00%) | 154 (1.63%) | 9,315 (98.37%) | 54 (35.06%) | 100 (64.94%) | 4,534 (48.67%) | 4,781 (51.33%) | Difference in birth year distribution in ARFID vs. controls (overall) | 14.27 | 0.003 |
| - 1997-2000 | 7,739 (100.00%) | 149 (1.93%) | 7,590 (98.07%) | 71 (47.65%) | 78 (52.35%) | 3,759 (49.53%) | 3,831 (50.47%) | | | |
| - 2001-2004 | 7,327 (100.00%) | 161 (2.20%) | 7,166 (97.80%) | 60 (37.27%) | 101 (62.73%) | 3,561 (49.69%) | 3,605 (50.31%) | | | |
| - 2005-2008 | 6,260 (100.00%) | 152 (2.43%) | 6,108 (97.57%) | 56 (36.84%) | 96 (63.16%) | 3,068 (50.23%) | 3,040 (49.77%) | | | |

Absolute number (n) and percentage of individuals with ARFID and controls are stated overall and per sex, age at CATSS assessment, and birth year. Row-wise percentages were calculated (separately for 1., 2., 3., and 4.). Birth years were pooled in groups of five (first group 1992–1996) or four (all other groups) consecutive years. Pearson $\chi^2$ tests were conducted to compare counts (highlighted with light-gray background) in ARFID vs. controls. *Abbreviations: ARFID, avoidant restrictive food intake disorder; CATSS, Child and Adolescent Twin Study in Sweden*.

**Table S2. Diagnostic classification of examined mental and somatic conditions in ARFID and controls**

| **ICD-chapters** | | | | | **Individual and grouped conditions within ICD-chapters** | | | | | | **Comments** |
|---|---|---|---|---|---|---|---|---|---|---|---|
| **Label** | **Description** | **ICD-10-codes** | **ICD-9-codes** | **Included as outcome?** | **Short label** | **Description** | **ICD-10-codes** | **ICD-9-codes** | **$n_{ARFID}$** | **Included as outcome?** | |
| **Cancer** | Malignant Neoplasms | C00-C97, D00-D09 | 140-209, 230-234 | Yes (malignant neoplasms and in situ neoplasms) | n/a | n/a | n/a | n/a | 13 | n/a | Chapter-level analysis; individual diagnoses were not analyzed (low prevalence) |
| **Endocrine Conditions** | Endocrine, nutritional, and metabolic conditions | E00-E90 | 240-279 | Yes | Cystic Fibrosis | Cystic fibrosis | E84 | 277.0 | <5 | No (low prevalence) | |
| | | | | | **Diabetes Mellitus** | Diabetes mellitus | E10, E11, E12, E13, E14 | 250 | 5 | Yes | |
| | | | | | **Lactose Intolerance** | Lactose intolerance | E73 | 271.3 | 11 | Yes | |
| | | | | | Lipid Metabolism | Disorders of lipoprotein and fatty acid metabolism | E71.3, E78 | 272 | <5 | No (low prevalence) | |
| | | | | | **Obesity** | Obesity | E66 | 278 | 7 | Yes | |
| | | | | | OtherFluid Homeostasis | Disturbances of fluid, electrolyte, acid-base balance (excl. volume depletion) | E87 | 276, excl. 276.5 | <5 | No (low prevalence) | |
| | | | | | **Pituitary Conditions** | Conditions of the pituitary gland | E23 | 253 | 9 | Yes | |
| | | | | | **Puberty Onset** | Disorders of puberty (delayed/precocious) | E30 | 259.0, 259.1 | 11 | Yes | |
| | | | | | **ShortStature** | Short stature | E34.3 | 259.4 | 28 | Yes | |
| | | | | | **Thyroid Conditions** | Thyroid conditions | E00, E01, E02, E03, E04, E05, E06, E07 | 240, 241, 242, 243, 244, 245, 246 | 7 | Yes | |
| | | | | | **Volume Depletion** | Volume depletion (abnormally decreased volume of circulating fluid [plasma]) | E86 | 276.5 | 24 | Yes | |
| **Mental Conditions** | Mental and behavioral conditions | F30-F39, F40-F42, F45, F50, F70-F79, F80-F89, F90-F98 | 296-299, 300, 307, 311-315, 317-319 | Yes (excl.: F00-F29 = organic mental disorders, psychoactive substance use, schizo-phrenia; F60 = adult personality and behavior) | **ADHD** | Attention deficit hyperactivity disorder | F90 | 314 | 108 | Yes | |
| | | | | | **Autism** | Autism (incl. childhood autism, atypical autism, Asperger syndrome, other pervasive developmental disorders) | F84.0, F84.1, F84.5, F84.8, F84.9 | 299.0 | 85 | Yes | |
| | | | | | Bipolar Disorder | Bipolar disorder | F30, F31 | 296 | <5 | No (low prevalence) | |
| | | | | | Bulimia Nervosa | Bulimia nervosa | F50.2, F50.3 | n/a | <5 | No (low prevalence) | Bulimia nervosa diagnosis according to ICD-9 (307.51) could not be ordered from Swedish Twin Registry (greatest level of detail |

| | | | | | | | | | | | |
|---|---|---|---|---|---|---|---|---|---|---|---|
| | | | | | | | | | | | provided was 307.5 [4 digits] = other and unspecified disorders of eating) |
| | | | | | **Conduct Disorder** | Conduct disorder | F91 | 312 | 9 | Yes | |
| | | | | | **Depression** | Depressive episode (single/recurrent), dysthymia | F32, F33, F34.1 | 296.2, 296.3, 300.4 | 16 | Yes | |
| | | | | | **Enuresis Encopresis** | Enuresis (nonorganic) | F98.0 | 307.6 | 17 | Yes | Related conditions, combined due to low prevalence |
| | | | | | | Encopresis (nonorganic) | F98.1 | 307.7 | | | |
| | | | | | **Intellectual Disability** | Intellectual disability | F70, F71, F72, F73, F74, F75, F76, F77, F78, F79 | 317, 318, 319 | 56 | Yes | |
| | | | | | **Motor Development** | Developmental disorders of motor function | F82 | 315.4 | 18 | Yes | |
| | | | | | **ObsessCompuls Disorder** | Obsessive-compulsive disorder | F42 | 300.3 | 5 | Yes | |
| | | | | | **OtherAnxiety Conditions** | Anxiety disorders excl. phobias (e.g., panic disorder, generalized anxiety disorder) | F41 | 300.0 | 20 | Yes | |
| | | | | | **OtherBehavior Emotion** | Other and unspecified behavioral and emotional disorders with onset in childhood and adolescence (e.g., Pica, impaired emotional/social functioning) | F93, F94, F98.3, F98.8, F98.9 | 313 | 14 | Yes | |
| | | | | | **Other Development** | Mixed specific, other, and unspecified developmental disorders (e.g., developmental agnosia) | F83, F88, F89 | 315.5, 315.8, 315.9 | 15 | Yes | |
| | | | | | **Phobias** | Phobic anxiety disorders (e.g., social phobia) | F40 | 300.2 | 5 | Yes | |
| | | | | | **Scholast Development** | Developmental disorders of scholastic skills (reading, spelling, arithmetical) | F81 | 315.0, 315.1, 315.2 | 13 | Yes | |
| | | | | | Somatoform Disorders | Somatoform disorders | F45 | 300.8 | <5 | No (low prevalence) | |
| | | | | | **Speech Development** | Developmental disorders of speech or language (incl. cluttering and stuttering) | F80, F98.5, F98.6 | 315.3 | 25 | Yes | |
| | | | | | **TicDisorders** | Tic disorders incl. Tourette syndrome | F95 | 307.2 | 14 | Yes | |
| Neurologic Conditions | Conditions of the nervous system | See condition-level for specification | See condition-level for specification | No (only selected conditions examined) | **CerebralPalsy** | Cerebral palsy and paralytic syndromes | G80, G81, G82, G83 | 342, 343, 344 | 25 | Yes | |
| | | | | | **Epilepsy** | Epilepsy | G40 | 345 | 36 | Yes | |
| **Circulatory Conditions** | Conditions of the circulatory system | I00-I99 | 390-459 | Yes | **Heart Pulmonary** | Heart and pulmonary heart conditions | I26, I27, I28, I30, I31, I32, I33, I34, I35, I36, I37, I38, I39, I40, I41, I42, | 415, 416, 417, 420, 421, 422, 423, 424, 425, | 12 | Yes | |

| | | | | | | | | | | | |
|---|---|---|---|---|---|---|---|---|---|---|---|
| | | | | | | | I43, I44, I45, I46, I47, I48, I49, I50, I51, I52 | 426, 427, 428, 429 | | | |
| | | | | | **VascArterial** | Cerebrovascular disorders, disorders of arteries/arterioles/capillaries, hypertension | I10, I11, I12, I13, I14, I15, I60, I61, I62, I63, I64, I65, I66, I67, I68, I69, I70, I71, I72, I73, I74, I75, I76, I77, I78, I79 | 401, 402, 403, 404, 405, 430, 431, 432, 433, 434, 435, 436, 437, 438, 440, 441, 442, 443, 444, 445, 446, 447, 448, 449 | 6 | Yes | |
| | | | | | **VascVenous Lymphatic** | Disorders of veins, lymphatic vessels, and lymph nodes | I80, I81, I82, I83, I84, I85, I86, I87, I88, I89 | 451, 452, 453, 454, 455, 456, 457, 458, 459 | 7 | Yes | |
| **Respiratory Conditions** | Conditions of the respiratory system | J00-J99 | 460-519 | Yes | **AcuteLower Respiratory** | Acute lower respiratory disorders (e.g., pneumonia, influenza, bronchitis) | J09, J10, J11, J12, J13, J14, J15, J16, J17, J18, J20, J21, J22 | 466, 480, 481, 482, 483, 484, 485, 486, 487, 488, 490 | 112 | Yes | |
| | | | | | **AcuteUpper Respiratory** | Acute upper respiratory infections (e.g., nasopharyngitis, non-streptococcal tonsillitis, laryngitis, and epiglottitis) | J00, J01, J02, J03, J04, J05, J06, excl. J03.0 | 460, 461, 462, 463, 464, 465 | 187 | Yes | Related conditions, combined for context |
| | | | | | | Streptococcal tonsillitis | J03.0, B95.0 | 034.0 | | | |
| | | | | | **Asthma** | Asthma | J45, J46 | 493 | 128 | Yes | |
| | | | | | **ChronicLower Respiratory** | Chronic lower respiratory disorders (e.g., chronic bronchitis, chronic obstructive pulmonary disorder) | J40, J41, J42, J43, J44, J47 | 491, 492, 494, 496 | 8 | Yes | |
| | | | | | **ChronicUpper Respiratory** | Chronic upper respiratory disorders (e.g., chronic sinusitis, chronic rhinitis, chronic nasopharyngitis, polyp, deviated nasal septum, disorders of vocal cords/larynx) | J31, J32, J33, J34, J37, J38, J39 | 470, 471, 472, 473, 476, 478 | 94 | Yes | Related conditions, combined due to low prevalence |
| | | | | | | Chronic disorders of tonsils and/or adenoids (e.g., inflammation, hypertrophy, peritonsillar abscess) | J35, J36 | 474, 475 | | | |
| | | | | | OtherRespiratory Conditions | Other respiratory conditions (e.g., affecting interstitial tissue and pleura, pulmonary edema, lung abscess, pyothorax, pneumothorax, emphysema, respiratory failure) | J80, J81, J82, J83, J84, J85, J86, J90, J91, J92, J93, J94, J95, J96, J97, J98, J99 | 510, 511, 512, 513, 514, 515, 516, 517, 518, 519 | 17 | No (combined/ grouped due to low prevalence, resulting group too general/ unspecific) | |
| | | | | | **Rhinitis** | Vasomotor and allergic rhinitis | J30 | 477 | 53 | Yes | |
| **Digestive Conditions** | Conditions of the digestive system | K00-K93 | 520-579 | Yes | **AnusRectum Conditions** | Conditions of anus/rectum (hemorrhage, hemorrhoids, fissure, fistula, abscess, prolapse) | K60, K61, K62, K64 | 565, 566, 569.0, 569.1, | 8 | Yes | |

| | | | | | | | | | | | |
|---|---|---|---|---|---|---|---|---|---|---|---|
| | | | | | | | | 569.2, 569.3, 569.4 | | | |
| | | | | | **Appendicitis** | Appendicitis | K35, K36, K37 | 540, 541, 542, 543 | 7 | Yes | |
| | | | | | BiliarySystem Conditions | Conditions of liver (e.g., hepatitis), gallbladder, biliary tract, pancreas | K70, K71, K72, K73, K74, K75, K76, K77, K83 | 570, 571, 572, 573, 574, 575, 576, 577 | <5 | No (low prevalence) | |
| | | | | | **CeliacDisease** | Celiac disease and intestinal malabsorption | K90 | 579 | 16 | Yes | |
| | | | | | **Constipation** | Constipation | K59.0 | 564.0 | 87 | Yes | |
| | | | | | **Dentofacial Anomalies** | Dentofacial anomalies (e.g., teeth [position], caries, embedded/impacted teeth, jaws) | K01, K02, K03, K07, K10 | 520, 521, 522, 523, 524, 525, 526 | 8 | Yes | |
| | | | | | **Diarrhea** | Functional diarrhea | K59.1 | 564.5 | 8 | Yes | |
| | | | | | **Dyspepsia** | Functional dyspepsia | K30 | 536.8 | 6 | Yes | |
| | | | | | **Gastrointestinal Bleeding** | Hematemesis, gastrointestinal hemorrhage, melena | K92.0, K92.1, K92.2 | 578 | 6 | Yes | |
| | | | | | **GERD** | Gastro-esophageal reflux disorder | K21 | 530.1, 530.8 | 32 | Yes | |
| | | | | | **Hernia** | Hernia (e.g., inguinal, umbilical, diaphragmatic) | K40, K41, K42, K43, K44, K45, K46 | 550, 551, 552, 553 | 40 | Yes | |
| | | | | | **IBS** | Irritable bowel syndrome and other/ unspecified functional intestinal conditions | K58, K59.8, K59.9 | 564.1, 564.8, 564.9 | 6 | Yes | |
| | | | | | **Ileus** | Ileus (paralytic) | K56 | 560.1 | 6 | Yes | |
| | | | | | **Noninfective IBD** | Crohn's disease and ulcerative colitis (Inflammatory Bowel Disease/IBD) | K50, K51 | 555, 556 | 33 | Yes | Related conditions, combined due to low prevalence |
| | | | | | | Noninfective (allergic/dietetic/food hypersensitivity) gastroenteritis/colitis excl. Crohn's disease or ulcerative colitis | K52.2, K52.8, K52.9 | 558.3 | | | |
| | | | | | **OtherUpper Gastrointest** | Conditions of esophagus, stomach, and duodenum (inflammation, ulcer, perforation, fistula, dyskinesia; excl. GERD) | K20, K22, K26, K29, K31 | 530, 531, 532, 533, 534, 535, 537, 538, excl. 530.1/530.8 | 19 | Yes | |
| | | | | | **Stomatitis** | Stomatitis and oral mucositis, oral mucosa lesions (incl. biting), oral cyst, disorders of tongue and salivary gland | K09, K11, K12, K13.1, K13.7, K14 | 527, 528, 529 | 16 | Yes | |
| Dermatologic Conditions | Conditions of the skin and subcutaneous tissue | See condition-level for specification | See condition-level for specification | No (only selected conditions examined) | **Bullous Dermatitis** | Bullous disorders incl. pemphigus/ pemphigoid | L10, L11, L12, L13, L14 | 694 | 7 | Yes | |
| | | | | | **OtherDermatitis Eczema** | Atopic dermatitis (incl. diaper/napkin dermatitis) | L20 | 691 | 68 | Yes | Related conditions, combined due to low prevalence |
| | | | | | | Seborrheic dermatitis | L21 | 690.1 | | | |

| | | | | | | | | | | | |
|---|---|---|---|---|---|---|---|---|---|---|---|
| | | | | | | Other/unspecified dermatitis/eczema (incl. infective dermatitis, dermatitis due to in-/ external substances, [allergic] contact dermatitis, diaper/napkin dermatitis) | L22, L23, L24, L25, L26, L27, L30 | 681.0, 692, 693 | | | |
| | | | | | | Lichen and Pruritis | L28, L29 | 697, 698 | | | |
| | | | | | **Psoriasis** | Psoriasis and other papulosquamous disorders | L40, L41, L42, L43, L44, L45 | 696 | 7 | Yes | |
| | | | | | **Urticaria** | Urticaria | L50 | 708 | 27 | Yes | |
| Musculo-skeletal Conditions | Conditions of the musculo-skeletal system and connective tissue | See condition-level for specification | See condition-level for specification | No (only selected conditions examined) | JuvenileArthritis | Juvenile arthritis | M08 | 714.3 | <5 | No (low prevalence) | |
| Genito-urinary Conditions | Conditions of the genitourinary system | See condition-level for specification | See condition-level for specification | No (only selected conditions examined) | Menstruation Problems | Impaired menstruation (irregularities, pain) | N91, N92, N93, N94 | 625, 626 | 5 | No (female only) | |
| | | | | | Nephrotic Syndrome | Nephrotic syndrome | N04 | 581 | <5 | No (low prevalence) | |
| **Perinatal Conditions** | Certain conditions originating in the perinatal period | P00-P96 | 760-779 | Yes | **FetalGrowth Retardation** | Conditions related to gestation length, fetal growth/malnutrition | P05, P07 | 764, 765 | 177 | Yes | |
| | | | | | **Maternal Complications** | Maternal complications (during pregnancy, labor, delivery) | P00, P01, P02, P03, P04 | 760, 761, 762, 763 | 48 | Yes | |
| | | | | | **Neonatal Jaundice** | Neonatal jaundice | P58, P59 | 774 | 91 | Yes | |
| | | | | | **PerinatCNS Conditions** | Perinatal intracranial hemorrhage (nontraumatic) and injury to central nervous system (traumatic) | P10, P11, P52 | 767.0, 767.4, 767.5, 767.7, 772.1, 772.2 | 16 | Yes | Related conditions, combined due to low prevalence |
| | | | | | | Convulsions, disturbances of cerebral status | P90, P91 | 779.0, 779.1, 779.2 | | | |
| | | | | | **PerinatEndo-crineMetabolic** | Perinatal endocrine and metabolic conditions (e.g., hypoglycemia) | P70, P71, P72, P73, P74 | 775 | 41 | Yes | |
| | | | | | **PerinatFeeding** | Feeding problems of the newborn | P92 | 779.3 | 21 | Yes | |
| | | | | | **Perinat Hematologic** | Perinatal hematological/hemolytic conditions (e.g., anemia) | P54, P55, P61 | 776 | 22 | Yes | |
| | | | | | **Perinat Infections** | Perinatal infections (e.g., sepsis, skin infections) | P35, P36, P37, P38, P39 | 771 | 29 | Yes | |
| | | | | | **PerinatRespirat Cardiovasc** | Perinatal respiratory and cardiovascular conditions (e.g., birth asphyxia, acute respiratory distress syndrome, pneumothorax, persistent fetal circulation) | P20, P21, P22, P23, P24, P25, P26, P27, P28, P29 | 768, 769, 770 | 75 | Yes | |
| **Congenital Conditions** | Congenital malformations, deformations, | Q00-Q99 | 740-759 | Yes | **Chromosomal Abnorm** | Chromosomal abnormalities | Q90, Q91, Q92, Q93, Q94, Q95, Q96, Q97, Q98, Q99 | 758 | 10 | Yes | |

| | | | | | | | | | | | |
|---|---|---|---|---|---|---|---|---|---|---|---|
| | chromosomal abnormalities | | | | **Circulatory SystemMalform** | Malformations of the circulatory system (e.g., atrial/ventricular septal defect, patent ductus arteriosus) | Q20, Q21, Q22, Q23, Q24, Q25, Q26, Q27, Q28 | 745, 746, 747 | 40 | Yes | |
| | | | | | **Digestive SystemMalform** | Cleft lip/palate | Q35, Q36, Q37 | 749 | 26 | Yes | Related conditions, combined due to low prevalence |
| | | | | | | Malformations of the digestive system (e.g., ankyloglossia) | Q38, Q39, Q40, Q41, Q42, Q43, Q44, Q45 | 750, 751 | | | |
| | | | | | **GenitalMalform** | Malformations of genitals | Q50, Q51, Q52, Q53, Q54, Q55, Q56 | 752 | 19 | Yes | |
| | | | | | **HeadMalform** | Malformations of eye, ear, face, and neck | Q10, Q11, Q12, Q13, Q14, Q15, Q16, Q17, Q18 | 743, 744 | 17 | Yes | |
| | | | | | **Musculoskeletal Malform** | Musculoskeletal malformations | Q65, Q66, Q67, Q68, Q69, Q70, Q71, Q72, Q73, Q74, Q75, Q76, Q77, Q78, Q79 | 754, 755, 756 | 44 | Yes | |
| | | | | | **NervousSystem Malform** | Malformations of the nervous system (e.g., hydrocephalus, spina bifida) | Q00, Q01, Q02, Q03, Q04, Q05, Q06, Q07 | 741, 742 | 20 | Yes | |
| | | | | | Respiratory SystemMalform | Malformations of the respiratory system | Q30, Q31, Q32, Q33, Q34 | 748 | <5 | No (low prevalence) | |
| | | | | | **UrinarySystem Malform** | Malformations of the urinary system | Q60, Q61, Q62, Q63, Q64 | 753 | 11 | Yes | |
| Symptoms | Symptoms, signs, and abnormal clinical and laboratory findings | See condition-level for specification | See condition-level for specification | No (only selected conditions examined) | **Dysphagia** | Dysphagia | R13 | 787.2 | 25 | Yes | |
| | | | | | **FebrileSeizures** | Febrile seizures/convulsions | R56.0 | 780.3 | 14 | Yes | |
| | | | | | **NauseaVomiting** | Nausea and vomiting | R11 | 787.0, 536.2 | 45 | Yes | |
| | | | | | Physiological Development | Lack of expected normal physiological development (delayed milestones, e.g., late talker, late walker) | R62.0, R62.9 | n/a | <5 | No (low prevalence) | |
| Trauma | Injury and other consequences of external causes | See condition-level for specification | See condition-level for specification | No (only selected conditions examined) | **Intracranial Injury** | Intracranial injury (e.g., concussion) | S06 | 850, 851, 852, 853, 854 | 41 | Yes | |

| | **Conditions grouped across ICD-chapters** | | | | | | Comments |
|---|---|---|---|---|---|---|---|
| | **Short label** | **Description** | **ICD-10-codes** | **ICD-9-codes** | $n_{ARFID}$ | **Included as outcome?** | |
| Multiple ICD-chapters | **Allergic Conditions** | Allergic rhinitis, asthma, allergic gastroenteritis, allergic contact dermatitis, urticaria | J30, J45, J46, K52.2, L23, L50 | 477, 493, 558.3, 692.9, 708 | 170 | Yes | Food allergies were not included (specific ICD-codes could not be ordered from |

| | | | | | | | |
|---|---|---|---|---|---|---|---|
| | | | | | | | Swedish Twin Registry) |
| | **Autoimmune Conditions** | Autoimmune thyroiditis, diabetes mellitus type I, Crohn's disease, ulcerous colitis, bullous skin conditions (incl. pemphigus/pemphigoid), atopic dermatitis, psoriasis, juvenile arthritis | E06.3, E10, K50, K51, L10, L11, L12, L13, L14, L20, L40, L41, L42, L43, L44, L45, M08 | 245.2, 250.0, 555, 556, 691, 694, 696, 3 | 58 | Yes | |
| | **PANS/PANDAS** | Pediatric acute-onset neuropsychiatric syndrome (PANS) including the PANS subgroup related to streptococcal infections (PANDAS) | B95.0, D89.8, D89.9, G96.8 | 041.0, 279.8, 279.9, 349.8 | <5 | No (low prevalence) | ICD-codes most used by clinicians comprise single codes from ICD-chapters B (infectious diseases), D (diseases of blood and blood-forming organs involving immune mechanism), and G (nervous system diseases) |

Analyzed/included conditions (ICD-chapters [1 digit], individual and grouped conditions within ICD-chapters [3-4 digits], grouped conditions across ICD-chapters [allergic and autoimmune conditions]) are highlighted in **bold**. Short labels of conditions within/across ICD-chapters are sorted in alphabetic order. Some conditions were combined/grouped for context or due to low prevalence (n<5, privacy reasons) of individual conditions (see comments column). *Abbreviations: ADHD, attention deficit hyperactivity disorder; ARFID, avoidant restrictive food intake disorder; CNS, central nervous system; GERD, gastro-esophageal reflux disorder; IBD, inflammatory bowel disease; IBS, irritable bowel syndrome; ICD, International Classification of Diseases.*

**Table S3. Prevalence of mental and somatic conditions in ARFID and controls overall, per sex, and per age group**

| **Condition** n (%) | **A. Overall** | | **B. Per sex** | | | | **C. Per age group** | | | | | |
|---|---|---|---|---|---|---|---|---|---|---|---|---|
| | **ARFID** | **Controls** | **ARFID** | | **Controls** | | **ARFID** | | | **Controls** | | |
| | | | **Female** | **Male** | **Female** | **Male** | **<6 years** | **6 to <12 years** | **12 to <18 years** | **<6 years** | **6 to <12 years** | **12 to <18 years** |
| ChapterE:EndocrineConditions | 109 (17.69%) | 2,119 (7.02%) | 52 (21.58%) | 57 (15.20%) | 1,039 (6.96%) | 1,080 (7.08%) | 56 (9.09%) | 40 (7.14%) | 13 (3.32%) | 853 (2.83%) | 746 (2.54%) | 520 (2.28%) |
| E:ShortStature | 28 (4.55%) | 423 (1.40%) | 18 (7.47%) | 10 (2.67%) | 183 (1.23%) | 240 (1.57%) | n/a | n/a | n/a | n/a | n/a | n/a |
| E:VolumeDepletion | 24 (3.90%) | 397 (1.32%) | 8 (3.32%) | 16 (4.27%) | 188 (1.26%) | 209 (1.37%) | n/a | n/a | n/a | n/a | n/a | n/a |
| E:Obesity | 7 (1.14%) | 351 (1.16%) | n/a | n/a | n/a | n/a | n/a | n/a | n/a | n/a | n/a | n/a |
| E:PubertyOnset | 11 (1.79%) | 224 (0.74%) | 6 (2.49%) | 5 (1.33%) | 111 (0.74%) | 113 (0.74%) | n/a | n/a | n/a | n/a | n/a | n/a |
| E:LactoseIntolerance | 11 (1.79%) | 214 (0.71%) | n/a | n/a | n/a | n/a | n/a | n/a | n/a | n/a | n/a | n/a |
| E:DiabetesMellitus | 5 (0.81%) | 190 (0.63%) | n/a | n/a | n/a | n/a | n/a | n/a | n/a | n/a | n/a | n/a |
| E:ThyroidConditions | 7 (1.14%) | 161 (0.53%) | n/a | n/a | n/a | n/a | n/a | n/a | n/a | n/a | n/a | n/a |
| E:PituitaryConditions | 9 (1.46%) | 76 (0.25%) | n/a | n/a | n/a | n/a | n/a | n/a | n/a | n/a | n/a | n/a |
| ChapterF:MentalConditions | 221 (35.88%) | 3,396 (11.25%) | 73 (30.29%) | 148 (39.47%) | 1,422 (9.53%) | 1,974 (12.94%) | 65 (10.55%) | 117 (21.23%) | 39 (11.78%) | 588 (1.95%) | 1,395 (4.71%) | 1,413 (6.27%) |
| F:ADHD | 108 (17.53%) | 1,168 (3.87%) | 33 (13.69%) | 75 (20.00%) | 356 (2.39%) | 812 (5.32%) | 9 (1.46%) | 70 (11.53%) | 29 (7.14%) | 39 (0.13%) | 556 (1.84%) | 573 (2.43%) |
| F:EnuresisEncopresis | 17 (2.76%) | 666 (2.21%) | n/a | n/a | n/a | n/a | n/a | n/a | n/a | n/a | n/a | n/a |
| F:OtherAnxietyConditions | 20 (3.25%) | 469 (1.55%) | 7 (2.90%) | 13 (3.47%) | 315 (2.11%) | 154 (1.01%) | n/a | n/a | n/a | n/a | n/a | n/a |
| F:Depression | 16 (2.60%) | 455 (1.51%) | n/a | n/a | n/a | n/a | n/a | n/a | n/a | n/a | n/a | n/a |
| F:Autism | 85 (13.80%) | 448 (1.48%) | 25 (10.37%) | 60 (16.00%) | 136 (0.91%) | 312 (2.04%) | 22 (3.57%) | 48 (8.08%) | 15 (3.65%) | 60 (0.20%) | 180 (0.60%) | 208 (0.87%) |
| F:SpeechDevelopment | 25 (4.06%) | 342 (1.13%) | 7 (2.90%) | 18 (4.80%) | 118 (0.79%) | 224 (1.47%) | n/a | n/a | n/a | n/a | n/a | n/a |
| F:IntellectualDisability | 56 (9.09%) | 282 (0.93%) | 16 (6.64%) | 40 (10.67%) | 94 (0.63%) | 188 (1.23%) | 22 (3.57%) | 25 (4.21%) | 9 (2.12%) | 63 (0.21%) | 129 (0.43%) | 90 (0.38%) |
| F:OtherBehaviorEmotion | 14 (2.27%) | 206 (0.68%) | n/a | n/a | n/a | n/a | n/a | n/a | n/a | n/a | n/a | n/a |
| F:ScholastDevelopment | 13 (2.11%) | 178 (0.59%) | n/a | n/a | n/a | n/a | n/a | n/a | n/a | n/a | n/a | n/a |
| F:ObsessCompulsDisorder | 5 (0.81%) | 149 (0.49%) | n/a | n/a | n/a | n/a | n/a | n/a | n/a | n/a | n/a | n/a |
| F:TicDisorders | 14 (2.27%) | 132 (0.44%) | n/a | n/a | n/a | n/a | n/a | n/a | n/a | n/a | n/a | n/a |
| F:ConductDisorder | 9 (1.46%) | 124 (0.41%) | n/a | n/a | n/a | n/a | n/a | n/a | n/a | n/a | n/a | n/a |
| F:MotorDevelopment | 18 (2.92%) | 110 (0.36%) | 9 (3.73%) | 9 (2.40%) | 38 (0.25%) | 72 (0.47%) | n/a | n/a | n/a | n/a | n/a | n/a |
| F:Phobias | 5 (0.81%) | 81 (0.27%) | n/a | n/a | n/a | n/a | n/a | n/a | n/a | n/a | n/a | n/a |
| F:OtherDevelopment | 15 (2.44%) | 57 (0.19%) | 5 (2.07%) | 10 (2.67%) | 25 (0.17%) | 32 (0.21%) | n/a | n/a | n/a | n/a | n/a | n/a |
| ChapterI:CirculatoryConditions | 25 (4.06%) | 639 (2.12%) | 6 (2.49%) | 19 (5.07%) | 270 (1.81%) | 369 (2.42%) | 9 (1.46%) | 11 (1.81%) | 5 (1.12%) | 185 (0.61%) | 213 (0.71%) | 241 (1.01%) |
| I:VascVenousLymphatic | 7 (1.14%) | 260 (0.86%) | n/a | n/a | n/a | n/a | n/a | n/a | n/a | n/a | n/a | n/a |
| I:HeartPulmonary | 12 (1.95%) | 205 (0.68%) | n/a | n/a | n/a | n/a | n/a | n/a | n/a | n/a | n/a | n/a |
| I:VascArterial | 6 (0.97%) | 148 (0.49%) | n/a | n/a | n/a | n/a | n/a | n/a | n/a | n/a | n/a | n/a |
| ChapterJ:RespiratoryConditions | 327 (53.08%) | 11,987 (39.72%) | 128 (53.11%) | 199 (53.07%) | 5,435 (36.42%) | 6,552 (42.94%) | 259 (42.05%) | 48 (13.45%) | 20 (8.26%) | 8,756 (29.01%) | 2,149 (10.03%) | 1,082 (6.72%) |
| J:AcuteUpperRespiratory | 187 (30.36%) | 6,139 (20.34%) | 72 (29.88%) | 115 (30.67%) | 2,796 (18.74%) | 3,343 (21.91%) | 152 (24.68%) | 25 (5.39%) | 10 (2.89%) | 5,002 (16.57%) | 761 (3.02%) | 376 (1.87%) |
| J:Asthma | 128 (20.78%) | 3,879 (12.85%) | 41 (17.01%) | 87 (23.20%) | 1,545 (10.35%) | 2,334 (15.30%) | 89 (14.45%) | 31 (5.88%) | 8 (2.12%) | 2,548 (8.44%) | 934 (3.38%) | 397 (1.85%) |
| J:ChronicUpperRespiratory | 94 (15.26%) | 3,570 (11.83%) | 44 (18.26%) | 50 (13.33%) | 1,599 (10.72%) | 1,971 (12.92%) | 58 (9.42%) | 27 (4.84%) | 9 (2.24%) | 1,903 (6.31%) | 1,123 (3.97%) | 544 (2.50%) |
| J:AcuteLowerRespiratory | 112 (18.18%) | 3,294 (10.91%) | 39 (16.18%) | 73 (19.47%) | 1,514 (10.15%) | 1,780 (11.67%) | n/a | n/a | n/a | n/a | n/a | n/a |
| J:Rhinitis | 53 (8.60%) | 1,709 (5.66%) | 21 (8.71%) | 32 (8.53%) | 659 (4.42%) | 1,050 (6.88%) | 8 (1.30%) | 30 (4.93%) | 15 (3.46%) | 273 (0.90%) | 901 (3.01%) | 535 (2.32%) |

| **Condition** n (%) | **A. Overall** | | **B. Per sex** | | | | **C. Per age group** | | | | | |
|---|---|---|---|---|---|---|---|---|---|---|---|---|
| | **ARFID** | **Controls** | **ARFID** | | **Controls** | | **ARFID** | | | **Controls** | | |
| | | | **Female** | **Male** | **Female** | **Male** | **<6 years** | **6 to <12 years** | **12 to <18 years** | **<6 years** | **6 to <12 years** | **12 to <18 years** |
| J:ChronicLowerRespiratory | 8 (1.30%) | 73 (0.24%) | n/a | n/a | n/a | n/a | n/a | n/a | n/a | n/a | n/a | n/a |
| ChapterK:DigestiveConditions | 210 (34.09%) | 5,462 (18.10%) | 80 (33.20%) | 130 (34.67%) | 2,430 (16.28%) | 3,032 (19.87%) | 123 (19.97%) | 62 (12.58%) | 25 (7.58%) | 2,992 (9.91%) | 1,499 (5.51%) | 971 (4.69%) |
| K:Constipation | 87 (14.12%) | 1,893 (6.27%) | 37 (15.35%) | 50 (13.33%) | 945 (6.33%) | 948 (6.21%) | 40 (6.49%) | 37 (6.42%) | 10 (2.42%) | 944 (3.13%) | 722 (2.47%) | 227 (0.99%) |
| K:Hernia | 40 (6.49%) | 1,260 (4.18%) | 6 (2.49%) | 34 (9.07%) | 258 (1.73%) | 1,002 (6.57%) | n/a | n/a | n/a | n/a | n/a | n/a |
| K:NoninfectiveIBD | 33 (5.36%) | 613 (2.03%) | 13 (5.39%) | 20 (5.33%) | 275 (1.84%) | 338 (2.22%) | 18 (2.92%) | 10 (1.67%) | 5 (1.13%) | 394 (1.31%) | 125 (0.42%) | 94 (0.40%) |
| K:Appendicitis | 7 (1.14%) | 517 (1.71%) | n/a | n/a | n/a | n/a | n/a | n/a | n/a | n/a | n/a | n/a |
| K:CeliacDisease | 16 (2.60%) | 393 (1.30%) | 7 (2.90%) | 9 (2.40%) | 255 (1.71%) | 138 (0.90%) | n/a | n/a | n/a | n/a | n/a | n/a |
| K:Stomatitis | 16 (2.60%) | 261 (0.86%) | 5 (2.07%) | 11 (2.93%) | 132 (0.88%) | 129 (0.85%) | n/a | n/a | n/a | n/a | n/a | n/a |
| K:DentofacialAnomalies | 8 (1.30%) | 247 (0.82%) | n/a | n/a | n/a | n/a | n/a | n/a | n/a | n/a | n/a | n/a |
| K:GERD | 32 (5.19%) | 223 (0.74%) | 13 (5.39%) | 19 (5.07%) | 108 (0.72%) | 115 (0.75%) | n/a | n/a | n/a | n/a | n/a | n/a |
| K:OtherUpperGastrointest | 19 (3.08%) | 206 (0.68%) | 8 (3.32%) | 11 (2.93%) | 118 (0.79%) | 88 (0.58%) | n/a | n/a | n/a | n/a | n/a | n/a |
| K:AnusRectumConditions | 8 (1.30%) | 191 (0.63%) | n/a | n/a | n/a | n/a | n/a | n/a | n/a | n/a | n/a | n/a |
| K:Diarrhea | 8 (1.30%) | 159 (0.53%) | n/a | n/a | n/a | n/a | n/a | n/a | n/a | n/a | n/a | n/a |
| K:IBS | 6 (0.97%) | 149 (0.49%) | n/a | n/a | n/a | n/a | n/a | n/a | n/a | n/a | n/a | n/a |
| K:GastrointestinalBleeding | 6 (0.97%) | 96 (0.32%) | n/a | n/a | n/a | n/a | n/a | n/a | n/a | n/a | n/a | n/a |
| K:Ileus | 6 (0.97%) | 66 (0.22%) | n/a | n/a | n/a | n/a | n/a | n/a | n/a | n/a | n/a | n/a |
| K:Dyspepsia | 6 (0.97%) | 58 (0.19%) | n/a | n/a | n/a | n/a | n/a | n/a | n/a | n/a | n/a | n/a |
| ChapterP:PerinatalConditions | 212 (34.42%) | 8,538 (28.29%) | 78 (32.37%) | 134 (35.73%) | 4,094 (27.44%) | 4,444 (29.13%) | n/a | n/a | n/a | n/a | n/a | n/a |
| P:FetalGrowthRetardation | 177 (28.73%) | 7,111 (23.56%) | 69 (28.63%) | 108 (28.80%) | 3,486 (23.36%) | 3,625 (23.76%) | n/a | n/a | n/a | n/a | n/a | n/a |
| P:NeonatalJaundice | 91 (14.77%) | 3,322 (11.01%) | 35 (14.52%) | 56 (14.93%) | 1,565 (10.49%) | 1,757 (11.52%) | n/a | n/a | n/a | n/a | n/a | n/a |
| P:PerinatRespiratCardiovasc | 75 (12.18%) | 2,787 (9.23%) | 26 (10.79%) | 49 (13.07%) | 1,216 (8.15%) | 1,571 (10.30%) | n/a | n/a | n/a | n/a | n/a | n/a |
| P:MaternalComplications | 48 (7.79%) | 2,225 (7.37%) | 17 (7.05%) | 31 (8.27%) | 1,104 (7.40%) | 1,121 (7.35%) | n/a | n/a | n/a | n/a | n/a | n/a |
| P:PerinatEndocrineMetabolic | 41 (6.66%) | 1,485 (4.92%) | 18 (7.47%) | 23 (6.13%) | 657 (4.40%) | 828 (5.43%) | n/a | n/a | n/a | n/a | n/a | n/a |
| P:PerinatInfections | 29 (4.71%) | 776 (2.57%) | 11 (4.56%) | 18 (4.80%) | 371 (2.49%) | 405 (2.65%) | n/a | n/a | n/a | n/a | n/a | n/a |
| P:PerinatHematologic | 22 (3.57%) | 769 (2.55%) | 7 (2.90%) | 15 (4.00%) | 370 (2.48%) | 399 (2.62%) | n/a | n/a | n/a | n/a | n/a | n/a |
| P:PerinatFeeding | 21 (3.41%) | 483 (1.60%) | 8 (3.32%) | 13 (3.47%) | 257 (1.72%) | 226 (1.48%) | n/a | n/a | n/a | n/a | n/a | n/a |
| P:PerinatCNSConditions | 16 (2.60%) | 188 (0.62%) | 6 (2.49%) | 10 (2.67%) | 80 (0.54%) | 108 (0.71%) | n/a | n/a | n/a | n/a | n/a | n/a |
| ChapterQ:CongenitalConditions | 138 (22.40%) | 3,608 (11.96%) | 57 (23.65%) | 81 (21.60%) | 1,470 (9.85%) | 2,138 (14.01%) | 108 (17.53%) | 17 (3.35%) | 13 (3.42%) | 2,614 (8.66%) | 629 (2.28%) | 365 (1.68%) |
| Q:MusculoskeletalMalform | 44 (7.14%) | 1,038 (3.44%) | 16 (6.64%) | 28 (7.47%) | 484 (3.24%) | 554 (3.63%) | 25 (4.06%) | 13 (2.20%) | 6 (1.37%) | 659 (2.18%) | 197 (0.67%) | 182 (0.78%) |
| Q:CirculatorySystemMalform | 40 (6.49%) | 843 (2.79%) | 21 (8.71%) | 19 (5.07%) | 451 (3.02%) | 392 (2.57%) | n/a | n/a | n/a | n/a | n/a | n/a |
| Q:GenitalMalform | 19 (3.08%) | 607 (2.01%) | n/a | n/a | n/a | n/a | n/a | n/a | n/a | n/a | n/a | n/a |
| Q:DigestiveSystemMalform | 26 (4.22%) | 491 (1.63%) | 11 (4.56%) | 15 (4.00%) | 158 (1.06%) | 333 (2.18%) | n/a | n/a | n/a | n/a | n/a | n/a |
| Q:HeadMalform | 17 (2.76%) | 409 (1.36%) | 7 (2.90%) | 10 (2.67%) | 183 (1.23%) | 226 (1.48%) | n/a | n/a | n/a | n/a | n/a | n/a |
| Q:UrinarySystemMalform | 11 (1.79%) | 184 (0.61%) | n/a | n/a | n/a | n/a | n/a | n/a | n/a | n/a | n/a | n/a |
| Q:NervousSystemMalform | 20 (3.25%) | 86 (0.28%) | 9 (3.73%) | 11 (2.93%) | 46 (0.31%) | 40 (0.26%) | n/a | n/a | n/a | n/a | n/a | n/a |
| Q:ChromosomalAbnorm | 10 (1.62%) | 52 (0.17%) | 5 (2.07%) | 5 (1.33%) | 23 (0.15%) | 29 (0.19%) | n/a | n/a | n/a | n/a | n/a | n/a |

| **Condition** n (%) | **A. Overall** | | **B. Per sex** | | | | **C. Per age group** | | | | | |
|---|---|---|---|---|---|---|---|---|---|---|---|---|
| | **ARFID** | **Controls** | **ARFID** | | **Controls** | | **ARFID** | | | **Controls** | | |
| | | | **Female** | **Male** | **Female** | **Male** | **<6 years** | **6 to <12 years** | **12 to <18 years** | **<6 years** | **6 to <12 years** | **12 to <18 years** |
| FurtherConditions | | | | | | | | | | | | |
| L:OtherDermatitisEczema | 68 (11.04%) | 2,284 (7.57%) | 28 (11.62%) | 40 (10.67%) | 1,142 (7.65%) | 1,142 (7.49%) | 41 (6.66%) | 12 (2.09%) | 15 (3.53%) | 1,202 (3.98%) | 725 (2.50%) | 357 (1.58%) |
| S:IntracranialInjury | 41 (6.66%) | 1,761 (5.84%) | 12 (4.98%) | 29 (7.73%) | 797 (5.34%) | 964 (6.32%) | 21 (3.41%) | 14 (2.35%) | 6 (1.38%) | 689 (2.28%) | 617 (2.09%) | 455 (1.98%) |
| L:Urticaria | 27 (4.38%) | 900 (2.98%) | 8 (3.32%) | 19 (5.07%) | 421 (2.82%) | 479 (3.14%) | n/a | n/a | n/a | n/a | n/a | n/a |
| R:NauseaVomiting | 45 (7.31%) | 649 (2.15%) | 18 (7.47%) | 27 (7.20%) | 320 (2.14%) | 329 (2.16%) | n/a | n/a | n/a | n/a | n/a | n/a |
| R:FebrileSeizures | 14 (2.27%) | 587 (1.95%) | 5 (2.07%) | 9 (2.40%) | 263 (1.76%) | 324 (2.12%) | n/a | n/a | n/a | n/a | n/a | n/a |
| G:Epilepsy | 36 (5.84%) | 318 (1.05%) | 8 (3.32%) | 28 (7.47%) | 150 (1.01%) | 168 (1.10%) | n/a | n/a | n/a | n/a | n/a | n/a |
| G:CerebralPalsy | 25 (4.06%) | 202 (0.67%) | 7 (2.90%) | 18 (4.80%) | 94 (0.63%) | 108 (0.71%) | n/a | n/a | n/a | n/a | n/a | n/a |
| L:Psoriasis | 7 (1.14%) | 186 (0.62%) | n/a | n/a | n/a | n/a | n/a | n/a | n/a | n/a | n/a | n/a |
| L:BullousDermatitis | 7 (1.14%) | 170 (0.56%) | n/a | n/a | n/a | n/a | n/a | n/a | n/a | n/a | n/a | n/a |
| R:Dysphagia | 25 (4.06%) | 120 (0.40%) | 6 (2.49%) | 19 (5.07%) | 63 (0.42%) | 57 (0.37%) | 12 (1.95%) | 7 (1.16%) | 6 (1.33%) | 47 (0.16%) | 48 (0.16%) | 25 (0.10%) |
| EntireChapterC:Cancer | 13 (2.11%) | 79 (0.26%) | 5 (2.07%) | 8 (2.13%) | 29 (0.19%) | 50 (0.33%) | n/a | n/a | n/a | n/a | n/a | n/a |
| GroupedConditions | | | | | | | | | | | | |
| AllergicConditions | 170 (27.60%) | 5,366 (17.78%) | 59 (24.48%) | 111 (29.60%) | 2,244 (15.04%) | 3,122 (20.46%) | 105 (17.05%) | 45 (8.81%) | 20 (5.59%) | 3,110 (10.31%) | 1,513 (5.59%) | 743 (3.60%) |
| AutoimmuneConditions | 58 (9.42%) | 2,218 (7.35%) | 28 (11.62%) | 30 (8.00%) | 1,087 (7.28%) | 1,131 (7.41%) | 34 (5.52%) | 12 (2.06%) | 12 (2.81%) | 1,133 (3.75%) | 689 (2.37%) | 396 (1.75%) |

Absolute number (n) and percentage of individuals with ARFID and controls are stated for each condition overall (A.), per sex (B.), and per age group (C.). Conditions are sorted in descending order of overall prevalence (A.) in controls within ICD-chapters, the group of further conditions, and grouped allergic and autoimmune conditions (highlighted with blue background). Conditions with n<5 individuals in ARFID or controls are not reported for privacy reasons (n/a). *Abbreviations: ADHD, attention deficit hyperactivity disorder; ARFID, avoidant restrictive food intake disorder; CNS, central nervous system; GERD, gastro-esophageal reflux disorder; IBD, inflammatory bowel disease; IBS, irritable bowel syndrome.*

**Table S4. Base Cox regression model estimates in ARFID vs. controls**

| Condition | HR | 95%CI LL | 95%CI UL | Robust SE | z statistic | p-value | Significant (*) after FDR/BH |
|---|---|---|---|---|---|---|---|
| ChapterE:EndocrineConditions | 2.73 | 2.21 | 3.36 | 0.11 | 9.45 | <0.0001 | * |
| E:PituitaryConditions | 5.56 | 2.74 | 11.30 | 0.36 | 4.75 | <0.0001 | * |
| E:ShortStature | 3.29 | 2.16 | 5.03 | 0.22 | 5.52 | <0.0001 | * |
| E:VolumeDepletion | 2.78 | 1.81 | 4.28 | 0.22 | 4.66 | <0.0001 | * |
| E:PubertyOnset | 2.65 | 1.38 | 5.09 | 0.33 | 2.92 | 0.003 | * |
| E:LactoseIntolerance | 2.52 | 1.30 | 4.89 | 0.34 | 2.74 | 0.006 | * |
| E:ThyroidConditions | 2.43 | 1.14 | 5.21 | 0.39 | 2.29 | 0.022 | * |
| E:DiabetesMellitus | 1.33 | 0.54 | 3.25 | 0.46 | 0.62 | 0.532 | |
| E:Obesity | 0.98 | 0.47 | 2.05 | 0.38 | -0.06 | 0.955 | |
| ChapterF:MentalConditions | 3.90 | 3.36 | 4.52 | 0.08 | 17.95 | <0.0001 | * |
| F:OtherDevelopment | 12.49 | 7.03 | 22.21 | 0.29 | 8.60 | <0.0001 | * |
| F:IntellectualDisability | 10.27 | 7.59 | 13.90 | 0.15 | 15.09 | <0.0001 | * |
| F:Autism | 9.68 | 7.52 | 12.47 | 0.13 | 17.60 | <0.0001 | * |
| F:MotorDevelopment | 7.41 | 4.33 | 12.68 | 0.27 | 7.30 | <0.0001 | * |
| F:ADHD | 4.86 | 3.93 | 6.00 | 0.11 | 14.69 | <0.0001 | * |
| F:TicDisorders | 4.80 | 2.64 | 8.71 | 0.30 | 5.15 | <0.0001 | * |
| F:ScholastDevelopment | 3.58 | 2.04 | 6.30 | 0.29 | 4.43 | <0.0001 | * |
| F:ConductDisorder | 3.58 | 1.84 | 6.96 | 0.34 | 3.76 | 0.0002 | * |
| F:Phobias | 3.57 | 1.46 | 8.74 | 0.46 | 2.78 | 0.005 | * |
| F:OtherBehaviorEmotion | 3.42 | 2.00 | 5.85 | 0.27 | 4.49 | <0.0001 | * |
| F:SpeechDevelopment | 3.32 | 2.18 | 5.05 | 0.21 | 5.60 | <0.0001 | * |
| F:OtherAnxietyConditions | 2.44 | 1.56 | 3.81 | 0.23 | 3.91 | <0.0001 | * |
| F:Depression | 2.13 | 1.28 | 3.52 | 0.26 | 2.93 | 0.003 | * |
| F:ObsessCompulsDisorder | 1.80 | 0.73 | 4.40 | 0.46 | 1.28 | 0.200 | |
| F:EnuresisEncopresis | 1.11 | 0.69 | 1.79 | 0.24 | 0.42 | 0.676 | |
| ChapterI:CirculatoryConditions | 1.96 | 1.29 | 2.97 | 0.21 | 3.17 | 0.002 | * |
| I:HeartPulmonary | 3.02 | 1.61 | 5.64 | 0.32 | 3.46 | 0.0005 | * |
| I:VascArterial | 2.02 | 0.89 | 4.60 | 0.42 | 1.68 | 0.094 | |
| I:VascVenousLymphatic | 1.29 | 0.61 | 2.73 | 0.38 | 0.66 | 0.512 | |
| ChapterJ:RespiratoryConditions | 1.42 | 1.26 | 1.60 | 0.06 | 5.72 | <0.0001 | * |
| J:ChronicLowerRespiratory | 4.87 | 2.35 | 10.10 | 0.37 | 4.25 | <0.0001 | * |
| J:AcuteLowerRespiratory | 1.59 | 1.30 | 1.94 | 0.10 | 4.57 | <0.0001 | * |
| J:Asthma | 1.57 | 1.30 | 1.88 | 0.09 | 4.74 | <0.0001 | * |
| J:Rhinitis | 1.52 | 1.14 | 2.01 | 0.14 | 2.89 | 0.004 | * |
| J:AcuteUpperRespiratory | 1.44 | 1.24 | 1.68 | 0.08 | 4.68 | <0.0001 | * |
| J:ChronicUpperRespiratory | 1.30 | 1.05 | 1.60 | 0.11 | 2.37 | 0.018 | * |
| ChapterK:DigestiveConditions | 2.03 | 1.76 | 2.35 | 0.07 | 9.58 | <0.0001 | * |
| K:GERD | 6.74 | 4.60 | 9.87 | 0.19 | 9.79 | <0.0001 | * |
| K:Dyspepsia | 5.67 | 2.44 | 13.16 | 0.43 | 4.04 | <0.0001 | * |
| K:OtherUpperGastrointest | 4.97 | 3.10 | 7.99 | 0.24 | 6.64 | <0.0001 | * |
| K:Ileus | 4.04 | 1.75 | 9.35 | 0.43 | 3.26 | 0.001 | * |
| K:Stomatitis | 3.02 | 1.78 | 5.13 | 0.27 | 4.09 | <0.0001 | * |
| K:GastrointestinalBleeding | 2.97 | 1.31 | 6.76 | 0.42 | 2.60 | 0.009 | * |
| K:NoninfectiveIBD | 2.58 | 1.76 | 3.76 | 0.19 | 4.90 | <0.0001 | * |
| K:Diarrhea | 2.34 | 1.14 | 4.79 | 0.37 | 2.32 | 0.020 | * |
| K:Constipation | 2.21 | 1.77 | 2.77 | 0.11 | 6.91 | <0.0001 | * |
| K:CeliacDisease | 2.11 | 1.28 | 3.47 | 0.26 | 2.92 | 0.004 | * |
| K:AnusRectumConditions | 2.04 | 1.00 | 4.15 | 0.36 | 1.97 | 0.049 | |
| K:IBS | 2.00 | 0.89 | 4.49 | 0.41 | 1.68 | 0.094 | |
| K:DentofacialAnomalies | 1.81 | 0.89 | 3.67 | 0.36 | 1.64 | 0.100 | |
| K:Hernia | 1.41 | 1.01 | 1.96 | 0.17 | 2.03 | 0.042 | |

| Condition | HR | 95%CI LL | 95%CI UL | Robust SE | z statistic | p-value | Significant (*) after FDR/BH |
|---|---|---|---|---|---|---|---|
| K:Appendicitis | 0.70 | 0.33 | 1.48 | 0.38 | -0.93 | 0.353 | |
| ChapterP:PerinatalConditions | 1.23 | 1.07 | 1.42 | 0.07 | 2.87 | 0.004 | * |
| P:PerinatCNSConditions | 3.99 | 2.37 | 6.71 | 0.26 | 5.23 | <0.0001 | * |
| P:PerinatFeeding | 2.04 | 1.29 | 3.23 | 0.23 | 3.07 | 0.002 | * |
| P:PerinatInfections | 1.80 | 1.22 | 2.67 | 0.20 | 2.96 | 0.003 | * |
| P:NeonatalJaundice | 1.35 | 1.09 | 1.67 | 0.11 | 2.76 | 0.006 | * |
| P:PerinatEndocrineMetabolic | 1.33 | 0.98 | 1.81 | 0.16 | 1.81 | 0.071 | |
| P:PerinatHematologic | 1.33 | 0.84 | 2.10 | 0.23 | 1.22 | 0.221 | |
| P:PerinatRespiratCardiovasc | 1.30 | 1.02 | 1.65 | 0.12 | 2.12 | 0.034 | * |
| P:FetalGrowthRetardation | 1.24 | 1.06 | 1.45 | 0.08 | 2.64 | 0.008 | * |
| P:MaternalComplications | 1.13 | 0.85 | 1.50 | 0.15 | 0.81 | 0.417 | |
| ChapterQ:CongenitalConditions | 1.89 | 1.58 | 2.26 | 0.09 | 7.00 | <0.0001 | * |
| Q:NervousSystemMalform | 11.50 | 7.06 | 18.75 | 0.25 | 9.80 | <0.0001 | * |
| Q:ChromosomalAbnorm | 9.21 | 4.68 | 18.13 | 0.35 | 6.42 | <0.0001 | * |
| Q:UrinarySystemMalform | 2.68 | 1.39 | 5.18 | 0.34 | 2.93 | 0.003 | * |
| Q:DigestiveSystemMalform | 2.40 | 1.62 | 3.57 | 0.20 | 4.34 | <0.0001 | * |
| Q:CirculatorySystemMalform | 2.35 | 1.70 | 3.26 | 0.17 | 5.13 | <0.0001 | * |
| Q:HeadMalform | 2.02 | 1.24 | 3.29 | 0.25 | 2.84 | 0.005 | * |
| Q:MusculoskeletalMalform | 2.01 | 1.48 | 2.73 | 0.16 | 4.43 | <0.0001 | * |
| Q:GenitalMalform | 1.28 | 0.81 | 2.03 | 0.23 | 1.07 | 0.286 | |
| FurtherConditions | | | | | | | |
| R:Dysphagia | 10.40 | 6.79 | 15.92 | 0.22 | 10.77 | <0.0001 | * |
| EntireChapterC:Cancer | 7.55 | 4.14 | 13.77 | 0.31 | 6.59 | <0.0001 | * |
| G:CerebralPalsy | 6.23 | 4.09 | 9.47 | 0.21 | 8.55 | <0.0001 | * |
| G:Epilepsy | 5.82 | 4.12 | 8.24 | 0.18 | 9.96 | <0.0001 | * |
| R:NauseaVomiting | 3.25 | 2.40 | 4.41 | 0.15 | 7.62 | <0.0001 | * |
| L:BullousDermatitis | 2.21 | 1.04 | 4.73 | 0.39 | 2.05 | 0.040 | |
| L:Psoriasis | 1.95 | 0.92 | 4.11 | 0.38 | 1.75 | 0.080 | |
| L:OtherDermatitisEczema | 1.46 | 1.13 | 1.88 | 0.13 | 2.92 | 0.004 | * |
| L:Urticaria | 1.44 | 0.97 | 2.13 | 0.20 | 1.80 | 0.072 | |
| S:IntracranialInjury | 1.17 | 0.86 | 1.60 | 0.16 | 0.99 | 0.323 | |
| R:FebrileSeizures | 1.13 | 0.67 | 1.92 | 0.27 | 0.45 | 0.651 | |
| GroupedConditions | | | | | | | |
| AllergicConditions | 1.56 | 1.32 | 1.83 | 0.08 | 5.34 | <0.0001 | * |
| AutoimmuneConditions | 1.29 | 0.98 | 1.70 | 0.14 | 1.79 | 0.073 | |

Stated are for each condition: hazard ratio in ARFID vs. controls (HR), lower/upper limits of the cluster-robust 95% confidence interval (95% CI LL/UL), cluster-robust standard error (SE), z statistic, raw p-value, and significance according to the false discovery rate-adjusted threshold (FDR/Benjamini-Hochberg procedure [BH]) from the base Cox regression model (predictor: group [ARFID vs. controls]; covariates: sex [female/male], birth year [1992–2008, factorized]; robust sandwich estimates given clustered twin data). Significantly different risks in ARFID vs. controls at $\alpha_{FDR}$=0.0343 are marked by an asterisk (*). Conditions are sorted in descending order of HRs within ICD-chapters, the group of further conditions, and grouped allergic and autoimmune conditions (highlighted with blue background). *Abbreviations: ADHD, attention deficit hyperactivity disorder; ARFID, avoidant restrictive food intake disorder; CNS, central nervous system; GERD, gastro-esophageal reflux disorder; IBD, inflammatory bowel disease; IBS, irritable bowel syndrome.*

**Table S5. Cumulative incidences of mental and somatic conditions before ages 6, 12, and 18 years in ARFID and controls**

| Condition | Age: <6 years | | | | | | Age: <12 years | | | | | | Age: <18 years | | | | | |
|---|---|---|---|---|---|---|---|---|---|---|---|---|---|---|---|---|---|---|
| | ARFID | | | Controls | | | ARFID | | | Controls | | | ARFID | | | Controls | | |
| | Cum. Inc. | 95%CI LL | 95%CI UL | Cum. Inc. | 95%CI LL | 95%CI UL | Cum. Inc. | 95%CI LL | 95%CI UL | Cum. Inc. | 95%CI LL | 95%CI UL | Cum. Inc. | 95%CI LL | 95%CI UL | Cum. Inc. | 95%CI LL | 95%CI UL |
| ChapterE:EndocrineConditions | 0.0925 | 0.0679 | 0.1165 | 0.0302 | 0.0258 | 0.0346 | 0.1604 | 0.1282 | 0.1915 | 0.0575 | 0.0514 | 0.0636 | 0.1993 | 0.1616 | 0.2353 | 0.0816 | 0.0736 | 0.0895 |
| E:PituitaryConditions | 0.0049 | 0.0000 | 0.0103 | 0.0005 | 0.0000 | 0.0010 | 0.0121 | 0.0031 | 0.0210 | 0.0023 | 0.0010 | 0.0035 | 0.0183 | 0.0059 | 0.0306 | 0.0023 | 0.0010 | 0.0035 |
| E:ShortStature | 0.0114 | 0.0018 | 0.0208 | 0.0041 | 0.0024 | 0.0057 | 0.0404 | 0.0222 | 0.0581 | 0.0102 | 0.0076 | 0.0129 | 0.0506 | 0.0300 | 0.0708 | 0.0161 | 0.0125 | 0.0197 |
| E:VolumeDepletion | 0.0292 | 0.0151 | 0.0431 | 0.0123 | 0.0095 | 0.0152 | 0.0398 | 0.0234 | 0.0559 | 0.0147 | 0.0115 | 0.0178 | 0.0398 | 0.0234 | 0.0559 | 0.0157 | 0.0123 | 0.0190 |
| E:PubertyOnset | 0.0049 | 0.0000 | 0.0119 | 0.0006 | 0.0000 | 0.0013 | 0.0102 | 0.0009 | 0.0194 | 0.0030 | 0.0014 | 0.0045 | 0.0263 | 0.0092 | 0.0431 | 0.0091 | 0.0061 | 0.0120 |
| E:LactoseIntolerance | 0.0097 | 0.0020 | 0.0175 | 0.0042 | 0.0025 | 0.0059 | 0.0186 | 0.0068 | 0.0303 | 0.0063 | 0.0042 | 0.0083 | 0.0186 | 0.0068 | 0.0303 | 0.0068 | 0.0047 | 0.0090 |
| E:ThyroidConditions | 0.0065 | 0.0001 | 0.0128 | 0.0013 | 0.0004 | 0.0022 | 0.0086 | 0.0010 | 0.0162 | 0.0029 | 0.0015 | 0.0043 | 0.0150 | 0.0030 | 0.0268 | 0.0060 | 0.0036 | 0.0084 |
| E:DiabetesMellitus | 0.0049 | 0.0000 | 0.0103 | 0.0016 | 0.0006 | 0.0026 | 0.0067 | 0.0001 | 0.0132 | 0.0037 | 0.0021 | 0.0053 | 0.0097 | 0.0009 | 0.0184 | 0.0073 | 0.0046 | 0.0099 |
| E:Obesity | 0.0016 | 0.0000 | 0.0048 | 0.0016 | 0.0005 | 0.0027 | 0.0104 | 0.0021 | 0.0187 | 0.0094 | 0.0067 | 0.0120 | 0.0126 | 0.0032 | 0.0220 | 0.0163 | 0.0124 | 0.0201 |
| ChapterF:MentalConditions | 0.1071 | 0.0814 | 0.1322 | 0.0235 | 0.0196 | 0.0275 | 0.3081 | 0.2674 | 0.3465 | 0.0854 | 0.0779 | 0.0929 | 0.4159 | 0.3669 | 0.4611 | 0.1549 | 0.1436 | 0.1661 |
| F:OtherDevelopment | 0.0146 | 0.0051 | 0.0240 | 0.0013 | 0.0004 | 0.0022 | 0.0237 | 0.0113 | 0.0359 | 0.0015 | 0.0005 | 0.0024 | 0.0259 | 0.0128 | 0.0388 | 0.0015 | 0.0005 | 0.0024 |
| F:IntellectualDisability | 0.0357 | 0.0203 | 0.0509 | 0.0024 | 0.0012 | 0.0037 | 0.0811 | 0.0576 | 0.1039 | 0.0076 | 0.0053 | 0.0098 | 0.1070 | 0.0770 | 0.1360 | 0.0135 | 0.0101 | 0.0168 |
| F:Autism | 0.0357 | 0.0197 | 0.0515 | 0.0021 | 0.0010 | 0.0033 | 0.1209 | 0.0912 | 0.1496 | 0.0090 | 0.0065 | 0.0115 | 0.1642 | 0.1281 | 0.1988 | 0.0193 | 0.0151 | 0.0234 |
| F:MotorDevelopment | 0.0162 | 0.0062 | 0.0262 | 0.0028 | 0.0013 | 0.0042 | 0.0285 | 0.0141 | 0.0427 | 0.0047 | 0.0028 | 0.0065 | 0.0309 | 0.0158 | 0.0457 | 0.0049 | 0.0030 | 0.0068 |
| F:ADHD | 0.0162 | 0.0062 | 0.0262 | 0.0024 | 0.0010 | 0.0038 | 0.1381 | 0.1078 | 0.1674 | 0.0301 | 0.0254 | 0.0347 | 0.2196 | 0.1780 | 0.2592 | 0.0617 | 0.0540 | 0.0693 |
| F:TicDisorders | 0.0016 | 0.0000 | 0.0048 | 0.0003 | 0.0000 | 0.0008 | 0.0173 | 0.0057 | 0.0287 | 0.0044 | 0.0026 | 0.0062 | 0.0293 | 0.0127 | 0.0456 | 0.0069 | 0.0044 | 0.0093 |
| F:ScholastDevelopment | 0.0016 | 0.0000 | 0.0048 | 0.0002 | 0.0000 | 0.0005 | 0.0113 | 0.0023 | 0.0203 | 0.0033 | 0.0018 | 0.0049 | 0.0335 | 0.0146 | 0.0521 | 0.0091 | 0.0062 | 0.0121 |
| F:ConductDisorder | 0.0049 | 0.0000 | 0.0103 | 0.0002 | 0.0000 | 0.0005 | 0.0137 | 0.0042 | 0.0231 | 0.0027 | 0.0013 | 0.0041 | 0.0169 | 0.0055 | 0.0281 | 0.0061 | 0.0036 | 0.0085 |
| F:Phobias | 0.0016 | 0.0000 | 0.0048 | 0.0003 | 0.0000 | 0.0008 | 0.0076 | 0.0001 | 0.0150 | 0.0007 | 0.0000 | 0.0015 | 0.0114 | 0.0008 | 0.0219 | 0.0030 | 0.0012 | 0.0049 |
| F:OtherBehaviorEmotion | 0.0049 | 0.0000 | 0.0103 | 0.0011 | 0.0003 | 0.0020 | 0.0188 | 0.0077 | 0.0297 | 0.0041 | 0.0023 | 0.0058 | 0.0290 | 0.0130 | 0.0448 | 0.0099 | 0.0068 | 0.0130 |
| F:SpeechDevelopment | 0.0227 | 0.0101 | 0.0352 | 0.0073 | 0.0050 | 0.0096 | 0.0350 | 0.0195 | 0.0502 | 0.0116 | 0.0088 | 0.0144 | 0.0450 | 0.0269 | 0.0628 | 0.0148 | 0.0115 | 0.0181 |
| F:OtherAnxietyConditions | 0.0016 | 0.0000 | 0.0048 | 0.0002 | 0.0000 | 0.0005 | 0.0227 | 0.0099 | 0.0353 | 0.0021 | 0.0008 | 0.0033 | 0.0509 | 0.0275 | 0.0736 | 0.0250 | 0.0195 | 0.0306 |
| F:Depression | 0.0000 | 0.0000 | 0.0000 | 0.0003 | 0.0000 | 0.0008 | 0.0115 | 0.0023 | 0.0206 | 0.0015 | 0.0005 | 0.0026 | 0.0417 | 0.0207 | 0.0623 | 0.0190 | 0.0142 | 0.0238 |
| F:ObsessCompulsDisorder | 0.0000 | 0.0000 | 0.0000 | 0.0002 | 0.0000 | 0.0005 | 0.0059 | 0.0000 | 0.0126 | 0.0017 | 0.0006 | 0.0028 | 0.0117 | 0.0012 | 0.0221 | 0.0088 | 0.0057 | 0.0119 |
| F:EnuresisEncopresis | 0.0032 | 0.0000 | 0.0077 | 0.0055 | 0.0037 | 0.0074 | 0.0265 | 0.0136 | 0.0393 | 0.0291 | 0.0246 | 0.0335 | 0.0294 | 0.0153 | 0.0434 | 0.0298 | 0.0253 | 0.0343 |
| ChapterI:CirculatoryConditions | 0.0146 | 0.0051 | 0.0240 | 0.0060 | 0.0041 | 0.0079 | 0.0346 | 0.0195 | 0.0495 | 0.0128 | 0.0099 | 0.0156 | 0.0519 | 0.0293 | 0.0740 | 0.0276 | 0.0226 | 0.0327 |
| I:HeartPulmonary | 0.0081 | 0.0010 | 0.0152 | 0.0028 | 0.0014 | 0.0041 | 0.0117 | 0.0030 | 0.0203 | 0.0045 | 0.0028 | 0.0063 | 0.0294 | 0.0084 | 0.0499 | 0.0086 | 0.0058 | 0.0114 |
| I:VascArterial | 0.0049 | 0.0000 | 0.0104 | 0.0011 | 0.0003 | 0.0020 | 0.0107 | 0.0021 | 0.0193 | 0.0017 | 0.0006 | 0.0027 | 0.0107 | 0.0021 | 0.0193 | 0.0042 | 0.0023 | 0.0062 |
| I:VascVenousLymphatic | 0.0032 | 0.0000 | 0.0077 | 0.0021 | 0.0010 | 0.0033 | 0.0102 | 0.0020 | 0.0184 | 0.0055 | 0.0036 | 0.0073 | 0.0135 | 0.0031 | 0.0238 | 0.0121 | 0.0087 | 0.0154 |
| ChapterJ:RespiratoryConditions | 0.4205 | 0.3786 | 0.4595 | 0.3258 | 0.3134 | 0.3380 | 0.5046 | 0.4613 | 0.5445 | 0.3960 | 0.3830 | 0.4087 | 0.5561 | 0.5107 | 0.5973 | 0.4509 | 0.4369 | 0.4645 |
| J:ChronicLowerRespiratory | 0.0114 | 0.0030 | 0.0197 | 0.0026 | 0.0013 | 0.0039 | 0.0130 | 0.0040 | 0.0219 | 0.0026 | 0.0013 | 0.0039 | 0.0130 | 0.0040 | 0.0219 | 0.0030 | 0.0015 | 0.0045 |
| J:AcuteLowerRespiratory | 0.1607 | 0.1302 | 0.1902 | 0.1120 | 0.1037 | 0.1203 | 0.1766 | 0.1442 | 0.2077 | 0.1231 | 0.1144 | 0.1316 | 0.1872 | 0.1535 | 0.2197 | 0.1305 | 0.1214 | 0.1395 |
| J:Asthma | 0.1461 | 0.1163 | 0.1749 | 0.1050 | 0.0969 | 0.1131 | 0.1996 | 0.1652 | 0.2325 | 0.1379 | 0.1287 | 0.1470 | 0.2240 | 0.1864 | 0.2600 | 0.1613 | 0.1511 | 0.1715 |

| Condition | Age: <6 years | | | | | | Age: <12 years | | | | | | Age: <18 years | | | | | |
|---|---|---|---|---|---|---|---|---|---|---|---|---|---|---|---|---|---|---|
| | ARFID | | | Controls | | | ARFID | | | Controls | | | ARFID | | | Controls | | |
| | Cum. Inc. | 95%CI LL | 95%CI UL | Cum. Inc. | 95%CI LL | 95%CI UL | Cum. Inc. | 95%CI LL | 95%CI UL | Cum. Inc. | 95%CI LL | 95%CI UL | Cum. Inc. | 95%CI LL | 95%CI UL | Cum. Inc. | 95%CI LL | 95%CI UL |
| J:Rhinitis | 0.0130 | 0.0040 | 0.0219 | 0.0115 | 0.0088 | 0.0143 | 0.0681 | 0.0462 | 0.0894 | 0.0472 | 0.0415 | 0.0528 | 0.1126 | 0.0805 | 0.1437 | 0.0794 | 0.0712 | 0.0875 |
| J:AcuteUpperRespiratory | 0.2484 | 0.2120 | 0.2831 | 0.1802 | 0.1701 | 0.1902 | 0.2903 | 0.2518 | 0.3269 | 0.2046 | 0.1940 | 0.2151 | 0.3175 | 0.2766 | 0.3560 | 0.2262 | 0.2147 | 0.2375 |
| J:ChronicUpperRespiratory | 0.0958 | 0.0715 | 0.1194 | 0.0722 | 0.0656 | 0.0789 | 0.1423 | 0.1123 | 0.1712 | 0.1102 | 0.1021 | 0.1182 | 0.1676 | 0.1343 | 0.1997 | 0.1412 | 0.1313 | 0.1510 |
| ChapterK:DigestiveConditions | 0.2013 | 0.1679 | 0.2334 | 0.1097 | 0.1016 | 0.1178 | 0.3081 | 0.2679 | 0.3462 | 0.1655 | 0.1557 | 0.1752 | 0.3754 | 0.3302 | 0.4175 | 0.2154 | 0.2035 | 0.2272 |
| K:GERD | 0.0308 | 0.0171 | 0.0444 | 0.0037 | 0.0021 | 0.0053 | 0.0470 | 0.0293 | 0.0645 | 0.0070 | 0.0047 | 0.0092 | 0.0582 | 0.0373 | 0.0787 | 0.0115 | 0.0083 | 0.0147 |
| K:Dyspepsia | 0.0016 | 0.0000 | 0.0048 | 0.0000 | 0.0000 | 0.0000 | 0.0098 | 0.0012 | 0.0183 | 0.0005 | 0.0000 | 0.0011 | 0.0124 | 0.0024 | 0.0223 | 0.0031 | 0.0012 | 0.0049 |
| K:OtherUpperGastrointest | 0.0146 | 0.0051 | 0.0240 | 0.0008 | 0.0001 | 0.0015 | 0.0220 | 0.0100 | 0.0338 | 0.0034 | 0.0019 | 0.0049 | 0.0385 | 0.0206 | 0.0560 | 0.0097 | 0.0065 | 0.0129 |
| K:Ileus | 0.0049 | 0.0000 | 0.0104 | 0.0024 | 0.0012 | 0.0037 | 0.0086 | 0.0010 | 0.0161 | 0.0024 | 0.0012 | 0.0037 | 0.0119 | 0.0019 | 0.0218 | 0.0028 | 0.0014 | 0.0042 |
| K:Stomatitis | 0.0162 | 0.0062 | 0.0261 | 0.0045 | 0.0029 | 0.0062 | 0.0243 | 0.0106 | 0.0378 | 0.0072 | 0.0050 | 0.0094 | 0.0295 | 0.0141 | 0.0446 | 0.0094 | 0.0067 | 0.0120 |
| K:GastrointestinalBleeding | 0.0081 | 0.0010 | 0.0152 | 0.0023 | 0.0011 | 0.0035 | 0.0081 | 0.0010 | 0.0152 | 0.0031 | 0.0017 | 0.0045 | 0.0105 | 0.0020 | 0.0189 | 0.0040 | 0.0023 | 0.0058 |
| K:NoninfectiveIBD | 0.0292 | 0.0144 | 0.0438 | 0.0156 | 0.0124 | 0.0188 | 0.0471 | 0.0282 | 0.0655 | 0.0195 | 0.0160 | 0.0231 | 0.0607 | 0.0385 | 0.0824 | 0.0253 | 0.0209 | 0.0297 |
| K:Diarrhea | 0.0097 | 0.0020 | 0.0174 | 0.0041 | 0.0024 | 0.0057 | 0.0115 | 0.0030 | 0.0199 | 0.0055 | 0.0036 | 0.0074 | 0.0137 | 0.0042 | 0.0231 | 0.0077 | 0.0051 | 0.0102 |
| K:Constipation | 0.0666 | 0.0457 | 0.0869 | 0.0343 | 0.0296 | 0.0389 | 0.1283 | 0.0995 | 0.1563 | 0.0628 | 0.0565 | 0.0690 | 0.1567 | 0.1232 | 0.1890 | 0.0761 | 0.0688 | 0.0833 |
| K:CeliacDisease | 0.0162 | 0.0062 | 0.0261 | 0.0070 | 0.0049 | 0.0091 | 0.0236 | 0.0113 | 0.0358 | 0.0111 | 0.0083 | 0.0138 | 0.0287 | 0.0145 | 0.0426 | 0.0133 | 0.0101 | 0.0166 |
| K:AnusRectumConditions | 0.0049 | 0.0000 | 0.0104 | 0.0028 | 0.0014 | 0.0041 | 0.0100 | 0.0020 | 0.0179 | 0.0046 | 0.0028 | 0.0063 | 0.0162 | 0.0043 | 0.0279 | 0.0065 | 0.0042 | 0.0087 |
| K:IBS | 0.0032 | 0.0000 | 0.0077 | 0.0029 | 0.0014 | 0.0044 | 0.0070 | 0.0001 | 0.0139 | 0.0040 | 0.0023 | 0.0057 | 0.0127 | 0.0022 | 0.0232 | 0.0072 | 0.0046 | 0.0098 |
| K:DentofacialAnomalies | 0.0016 | 0.0000 | 0.0048 | 0.0006 | 0.0000 | 0.0013 | 0.0054 | 0.0000 | 0.0115 | 0.0029 | 0.0015 | 0.0043 | 0.0197 | 0.0055 | 0.0338 | 0.0133 | 0.0095 | 0.0171 |
| K:Hernia | 0.0617 | 0.0420 | 0.0810 | 0.0390 | 0.0340 | 0.0439 | 0.0650 | 0.0443 | 0.0852 | 0.0447 | 0.0394 | 0.0500 | 0.0650 | 0.0443 | 0.0852 | 0.0483 | 0.0426 | 0.0540 |
| K:Appendicitis | 0.0032 | 0.0000 | 0.0077 | 0.0015 | 0.0005 | 0.0024 | 0.0068 | 0.0001 | 0.0135 | 0.0097 | 0.0071 | 0.0123 | 0.0157 | 0.0035 | 0.0277 | 0.0219 | 0.0173 | 0.0266 |
| ChapterP:PerinatalConditions | n/a | n/a | n/a | n/a | n/a | n/a | n/a | n/a | n/a | n/a | n/a | n/a | n/a | n/a | n/a | n/a | n/a | n/a |
| ChapterQ:CongenitalConditions | 0.1769 | 0.1452 | 0.2075 | 0.0998 | 0.0922 | 0.1074 | 0.2047 | 0.1710 | 0.2371 | 0.1217 | 0.1133 | 0.1300 | 0.2428 | 0.2042 | 0.2795 | 0.1369 | 0.1278 | 0.1460 |
| Q:NervousSystemMalform | 0.0325 | 0.0184 | 0.0464 | 0.0024 | 0.0012 | 0.0037 | 0.0325 | 0.0184 | 0.0464 | 0.0028 | 0.0014 | 0.0041 | 0.0325 | 0.0184 | 0.0464 | 0.0030 | 0.0016 | 0.0044 |
| Q:ChromosomalAbnorm | 0.0114 | 0.0030 | 0.0197 | 0.0013 | 0.0004 | 0.0022 | 0.0130 | 0.0040 | 0.0219 | 0.0013 | 0.0004 | 0.0022 | 0.0207 | 0.0067 | 0.0345 | 0.0013 | 0.0004 | 0.0022 |
| Q:UrinarySystemMalform | 0.0146 | 0.0041 | 0.0250 | 0.0055 | 0.0036 | 0.0074 | 0.0166 | 0.0054 | 0.0277 | 0.0063 | 0.0043 | 0.0084 | 0.0199 | 0.0070 | 0.0327 | 0.0063 | 0.0043 | 0.0084 |
| Q:DigestiveSystemMalform | 0.0390 | 0.0236 | 0.0541 | 0.0166 | 0.0133 | 0.0198 | 0.0390 | 0.0236 | 0.0541 | 0.0185 | 0.0151 | 0.0219 | 0.0441 | 0.0272 | 0.0608 | 0.0196 | 0.0160 | 0.0231 |
| Q:CirculatorySystemMalform | 0.0584 | 0.0392 | 0.0773 | 0.0265 | 0.0223 | 0.0306 | 0.0633 | 0.0434 | 0.0828 | 0.0279 | 0.0236 | 0.0322 | 0.0663 | 0.0455 | 0.0866 | 0.0296 | 0.0251 | 0.0340 |
| Q:HeadMalform | 0.0195 | 0.0085 | 0.0303 | 0.0076 | 0.0055 | 0.0098 | 0.0264 | 0.0135 | 0.0390 | 0.0125 | 0.0097 | 0.0154 | 0.0289 | 0.0152 | 0.0424 | 0.0136 | 0.0106 | 0.0166 |
| Q:MusculoskeletalMalform | 0.0422 | 0.0256 | 0.0586 | 0.0260 | 0.0219 | 0.0300 | 0.0637 | 0.0434 | 0.0837 | 0.0327 | 0.0282 | 0.0372 | 0.0805 | 0.0563 | 0.1042 | 0.0419 | 0.0364 | 0.0474 |
| Q:GenitalMalform | 0.0292 | 0.0159 | 0.0424 | 0.0211 | 0.0175 | 0.0247 | 0.0309 | 0.0172 | 0.0444 | 0.0269 | 0.0227 | 0.0310 | 0.0309 | 0.0172 | 0.0444 | 0.0276 | 0.0234 | 0.0318 |
| FurtherConditions | | | | | | | | | | | | | | | | | | |
| R:Dysphagia | 0.0195 | 0.0085 | 0.0303 | 0.0011 | 0.0003 | 0.0020 | 0.0320 | 0.0177 | 0.0461 | 0.0038 | 0.0022 | 0.0054 | 0.0485 | 0.0291 | 0.0675 | 0.0050 | 0.0030 | 0.0069 |
| EntireChapterC:Cancer | 0.0114 | 0.0030 | 0.0197 | 0.0018 | 0.0007 | 0.0028 | 0.0168 | 0.0064 | 0.0271 | 0.0027 | 0.0014 | 0.0040 | 0.0247 | 0.0110 | 0.0382 | 0.0031 | 0.0016 | 0.0046 |
| G:CerebralPalsy | 0.0406 | 0.0249 | 0.0560 | 0.0044 | 0.0027 | 0.0060 | 0.0406 | 0.0249 | 0.0560 | 0.0052 | 0.0034 | 0.0070 | 0.0406 | 0.0249 | 0.0560 | 0.0054 | 0.0036 | 0.0073 |
| G:Epilepsy | 0.0373 | 0.0223 | 0.0522 | 0.0044 | 0.0027 | 0.0060 | 0.0535 | 0.0352 | 0.0713 | 0.0098 | 0.0072 | 0.0123 | 0.0640 | 0.0431 | 0.0844 | 0.0127 | 0.0096 | 0.0159 |

| Condition | Age: <6 years | | | | | | Age: <12 years | | | | | | Age: <18 years | | | | | |
|---|---|---|---|---|---|---|---|---|---|---|---|---|---|---|---|---|---|---|
| | ARFID | | | Controls | | | ARFID | | | ARFID | | | Controls | | | ARFID | | |
| | Cum. Inc. | 95%CI LL | 95%CI UL | Cum. Inc. | 95%CI LL | 95%CI UL | Cum. Inc. | 95%CI LL | 95%CI UL | Cum. Inc. | 95%CI LL | 95%CI UL | Cum. Inc. | 95%CI LL | 95%CI UL | Cum. Inc. | 95%CI LL | 95%CI UL |
| R:NauseaVomiting | 0.0503 | 0.0329 | 0.0675 | 0.0131 | 0.0103 | 0.0160 | 0.0680 | 0.0476 | 0.0880 | 0.0197 | 0.0162 | 0.0233 | 0.0786 | 0.0558 | 0.1008 | 0.0247 | 0.0205 | 0.0290 |
| L:BullousDermatitis | 0.0114 | 0.0030 | 0.0197 | 0.0047 | 0.0029 | 0.0065 | 0.0114 | 0.0030 | 0.0197 | 0.0052 | 0.0033 | 0.0070 | 0.0114 | 0.0030 | 0.0197 | 0.0054 | 0.0035 | 0.0073 |
| L:Psoriasis | 0.0049 | 0.0000 | 0.0104 | 0.0015 | 0.0005 | 0.0024 | 0.0083 | 0.0010 | 0.0154 | 0.0047 | 0.0029 | 0.0065 | 0.0133 | 0.0033 | 0.0232 | 0.0093 | 0.0064 | 0.0122 |
| L:OtherDermatitisEczema | 0.0666 | 0.0454 | 0.0872 | 0.0464 | 0.0411 | 0.0518 | 0.0879 | 0.0639 | 0.1113 | 0.0759 | 0.0689 | 0.0828 | 0.1268 | 0.0962 | 0.1564 | 0.0964 | 0.0880 | 0.1047 |
| L:Urticaria | 0.0244 | 0.0114 | 0.0372 | 0.0148 | 0.0118 | 0.0178 | 0.0292 | 0.0152 | 0.0431 | 0.0255 | 0.0215 | 0.0295 | 0.0594 | 0.0351 | 0.0830 | 0.0338 | 0.0287 | 0.0389 |
| S:IntracranialInjury | 0.0357 | 0.0210 | 0.0502 | 0.0266 | 0.0226 | 0.0306 | 0.0586 | 0.0395 | 0.0773 | 0.0465 | 0.0411 | 0.0519 | 0.0753 | 0.0521 | 0.0980 | 0.0693 | 0.0619 | 0.0767 |
| R:FebrileSeizures | 0.0195 | 0.0085 | 0.0303 | 0.0185 | 0.0150 | 0.0220 | 0.0213 | 0.0097 | 0.0327 | 0.0187 | 0.0152 | 0.0222 | 0.0243 | 0.0114 | 0.0370 | 0.0193 | 0.0157 | 0.0229 |
| GroupedConditions | | | | | | | | | | | | | | | | | | |
| AllergicConditions | 0.1721 | 0.1402 | 0.2028 | 0.1252 | 0.1165 | 0.1338 | 0.2496 | 0.2121 | 0.2854 | 0.1775 | 0.1674 | 0.1875 | 0.3071 | 0.2633 | 0.3482 | 0.2217 | 0.2098 | 0.2334 |
| AutoimmuneConditions | 0.0568 | 0.0374 | 0.0758 | 0.0433 | 0.0381 | 0.0485 | 0.0758 | 0.0533 | 0.0977 | 0.0707 | 0.0640 | 0.0774 | 0.1063 | 0.0779 | 0.1338 | 0.0922 | 0.0839 | 0.1004 |

Cumulative incidence (Cum. Inc., absolute risk) as the probability (proportion) of having any of the analyzed conditions before three consecutive time points (i.e., before 6, 12, and 18 years of age) is stated in ARFID and controls, including lower/upper limits of the cluster-robust 95% confidence interval (95% CI LL/UL). Perinatal conditions were excluded [n/a] given their occurrence during the perinatal period and lack of age-related dynamics). Cumulative incidence was estimated using the Kaplan-Meier method (Cum. Inc. = 1 - Kaplan-Meier survival estimate, accounting for censoring) after creating a modified analysis sample by matching n=10 unexposed individuals to each (n=1) individual twin with ARFID, stratified by sex and birth year. Conditions are sorted in accordance with Table S4, i.e., in descending order of base cox regression model hazard ratios within ICD-chapters, the group of further conditions, and grouped allergic and autoimmune conditions (highlighted with blue background). *Abbreviations: ADHD, attention deficit hyperactivity disorder; ARFID, avoidant restrictive food intake disorder; CNS, central nervous system; GERD, gastro-esophageal reflux disorder; IBD, inflammatory bowel disease; IBS, irritable bowel syndrome.*

**Table S6. Number of distinct overall, all mental, and all somatic diagnoses in ARFID vs. controls**

| Condition | ARFID | | Controls | | Poisson regression model statistics | | | | | | |
|---|---|---|---|---|---|---|---|---|---|---|---|
| | Mean | SEM | Mean | SEM | IRR | 95%CI LL | 95%CI UL | Robust SE | z statistic | p-value | Significant (*) after FDR/BH |
| All diagnoses | 6.27 | 0.28 | 2.78 | 0.02 | 2.18 | 1.99 | 2.39 | 0.05 | 16.59 | <0.0001 | * |
| Mental diagnoses | 1.01 | 0.07 | 0.22 | 0.00 | 4.65 | 3.99 | 5.42 | 0.08 | 19.73 | <0.0001 | * |
| Somatic diagnoses | 5.26 | 0.26 | 2.56 | 0.02 | 1.98 | 1.79 | 2.19 | 0.05 | 13.55 | <0.0001 | * |

Mean and standard error of the mean (SEM) for number of *all* distinct diagnoses (i.e., unique diagnostic ICD-codes), number of distinct *mental* diagnoses, and number of distinct *somatic* diagnoses per individual twin are given in ARFID and controls. As test statistics, incidence rate ratio in ARFID vs. controls (IRR), lower/upper limits of the cluster-robust 95% confidence interval (95% CI LL/UL), cluster-robust standard error (SE), z statistic, raw p-value, and significance according to false discovery rate-adjusted threshold (FDR/Benjamini-Hochberg procedure [BH]) are stated for all distinct diagnoses, distinct mental diagnoses, and distinct somatic diagnoses. Statistics were obtained with Poisson regression models (predictor: group [ARFID vs. controls]; covariates: sex [female/male], birth year [1992–2008, factorized]; robust sandwich estimates given clustered twin data; offset term to account for follow-up/exposure time). Significantly different incidence rates in ARFID vs. controls at $\alpha_{FDR}$=0.0343 are marked by an asterisk (*). *Abbreviation: ARFID, avoidant restrictive food intake disorder.*

**Table S7. Number of inpatient days due to any, any mental, and any somatic diagnosis in ARFID vs. controls**

| Condition | ARFID | | Controls | | Poisson regression model statistics | | | | | | |
|---|---|---|---|---|---|---|---|---|---|---|---|
| | Mean | SEM | Mean | SEM | IRR | 95%CI LL | 95%CI UL | Robust SE | z statistic | p-value | Significant (*) after FDR/BH |
| Any diagnosis | 26.61 | 2.73 | 9.18 | 0.13 | 2.88 | 2.34 | 3.55 | 0.11 | 9.96 | <0.0001 | * |
| Mental diagnosis | 1.00 | 0.58 | 0.24 | 0.04 | 5.50 | 1.72 | 17.60 | 0.59 | 2.87 | 0.004 | * |
| Somatic diagnosis | 23.03 | 2.44 | 8.49 | 0.12 | 2.68 | 2.16 | 3.32 | 0.11 | 8.97 | <0.0001 | * |

Mean and standard error of the mean (SEM) for number of inpatient days per individual twin due to any diagnosis (i.e., unique diagnostic ICD-code), any mental diagnosis (as main diagnosis/cause of hospitalization), and any somatic diagnosis (as main diagnosis/cause of hospitalization) are given in ARFID and controls. As test statistics, incidence rate ratio in ARFID vs. controls (IRR), lower/upper limits of the cluster-robust 95% confidence interval (95% CI LL/UL), cluster-robust standard error (SE), z statistic, raw p-value, and significance according to false discovery rate-adjusted threshold (FDR/Benjamini-Hochberg procedure [BH]) are stated for any diagnosis, any mental diagnosis, and any somatic diagnosis. Statistics were obtained with Poisson regression models (predictor: group [ARFID vs. controls]; covariates: sex [female/male], birth year [1992–2008, factorized]; robust sandwich estimates given clustered twin data; offset term to account for follow-up/exposure time). Significantly different incidence rates in ARFID vs. controls at $\alpha_{FDR}$=0.0343 are marked by an asterisk (*). *Abbreviation: ARFID, avoidant restrictive food intake disorder.*

**Table S8. Sex-stratified Cox regression model estimates in ARFID vs. controls**

| Condition | Sex: female | | | | Sex: male | | | | Comparison female vs. male | | | | |
|---|---|---|---|---|---|---|---|---|---|---|---|---|---|
| | $HR_{female}$ | 95%CI LL | 95%CI UL | p-value | $HR_{male}$ | 95%CI LL | 95%CI UL | p-value | $HR_{female}$/ $HR_{male}$ | 95%CI LL | 95%CI UL | p-value | Significant (*) after FDR/BH |
| E:ShortStature | 6.10 | 3.56 | 10.44 | <0.0001 | 1.83 | 0.92 | 3.62 | 0.085 | 3.50 | 1.47 | 8.33 | 0.005 | |
| F:MotorDevelopment | 14.10 | 6.62 | 30.00 | <0.0001 | 4.90 | 2.29 | 10.48 | <0.0001 | 3.16 | 1.07 | 9.29 | 0.037 | |
| Q:DigestiveSystemMalform | 4.24 | 2.30 | 7.78 | <0.0001 | 1.84 | 1.10 | 3.06 | 0.020 | 2.38 | 1.07 | 5.27 | 0.033 | |
| Q:ChromosomalAbnorm | 12.95 | 4.89 | 34.29 | <0.0001 | 6.84 | 2.71 | 17.27 | <0.0001 | 1.89 | 0.49 | 7.27 | 0.353 | |
| E:PubertyOnset | 3.59 | 1.40 | 9.19 | 0.008 | 2.20 | 0.91 | 5.32 | 0.081 | 1.81 | 0.50 | 6.62 | 0.368 | |
| J:ChronicUpperRespiratory | 1.77 | 1.30 | 2.40 | 0.0003 | 1.04 | 0.78 | 1.39 | 0.790 | 1.72 | 1.13 | 2.62 | 0.011 | |
| ChapterQ:CongenitalConditions | 2.52 | 1.93 | 3.30 | <0.0001 | 1.61 | 1.27 | 2.03 | <0.0001 | 1.60 | 1.12 | 2.27 | 0.009 | |
| J:Rhinitis | 2.03 | 1.31 | 3.13 | 0.001 | 1.30 | 0.90 | 1.88 | 0.170 | 1.59 | 0.90 | 2.82 | 0.110 | |
| P:PerinatEndocrineMetabolic | 1.70 | 1.06 | 2.72 | 0.026 | 1.13 | 0.75 | 1.70 | 0.569 | 1.54 | 0.82 | 2.87 | 0.177 | |
| ChapterE:EndocrineConditions | 3.44 | 2.50 | 4.72 | <0.0001 | 2.30 | 1.75 | 3.02 | <0.0001 | 1.52 | 1.00 | 2.31 | 0.051 | |
| Q:CirculatorySystemMalform | 2.93 | 1.89 | 4.54 | <0.0001 | 1.92 | 1.18 | 3.12 | 0.008 | 1.51 | 0.79 | 2.91 | 0.213 | |
| EntireChapterC:Cancer | 10.50 | 3.93 | 28.05 | <0.0001 | 6.67 | 3.15 | 14.13 | <0.0001 | 1.51 | 0.44 | 5.21 | 0.515 | |
| AutoimmuneConditions | 1.64 | 1.11 | 2.41 | 0.013 | 1.09 | 0.74 | 1.60 | 0.677 | 1.51 | 0.88 | 2.59 | 0.138 | |
| F:Autism | 12.10 | 7.59 | 19.29 | <0.0001 | 8.97 | 6.66 | 12.09 | <0.0001 | 1.42 | 0.81 | 2.48 | 0.215 | |
| F:ADHD | 5.94 | 4.05 | 8.72 | <0.0001 | 4.47 | 3.46 | 5.76 | <0.0001 | 1.40 | 0.88 | 2.22 | 0.157 | |
| Q:HeadMalform | 2.27 | 1.06 | 4.88 | 0.035 | 1.87 | 0.99 | 3.52 | 0.053 | 1.27 | 0.47 | 3.44 | 0.638 | |
| ChapterJ:RespiratoryConditions | 1.60 | 1.33 | 1.93 | <0.0001 | 1.32 | 1.13 | 1.54 | 0.0004 | 1.21 | 0.95 | 1.55 | 0.116 | |
| P:PerinatCNSConditions | 4.35 | 1.89 | 9.99 | 0.0005 | 3.83 | 1.97 | 7.46 | <0.0001 | 1.20 | 0.41 | 3.53 | 0.734 | |
| K:NoninfectiveIBD | 2.88 | 1.65 | 5.04 | 0.0002 | 2.45 | 1.49 | 4.03 | 0.0004 | 1.19 | 0.57 | 2.48 | 0.649 | |
| ChapterK:DigestiveConditions | 2.24 | 1.79 | 2.80 | <0.0001 | 1.92 | 1.60 | 2.32 | <0.0001 | 1.18 | 0.89 | 1.57 | 0.260 | |
| K:Constipation | 2.41 | 1.73 | 3.37 | <0.0001 | 2.09 | 1.55 | 2.80 | <0.0001 | 1.16 | 0.75 | 1.80 | 0.501 | |
| F:IntellectualDisability | 10.48 | 6.01 | 18.26 | <0.0001 | 10.09 | 7.06 | 14.42 | <0.0001 | 1.14 | 0.58 | 2.23 | 0.702 | |
| J:AcuteUpperRespiratory | 1.57 | 1.23 | 2.00 | 0.0003 | 1.38 | 1.13 | 1.68 | 0.001 | 1.14 | 0.83 | 1.56 | 0.409 | |
| K:GERD | 7.43 | 4.17 | 13.24 | <0.0001 | 6.57 | 3.96 | 10.91 | <0.0001 | 1.12 | 0.52 | 2.40 | 0.780 | |
| AllergicConditions | 1.68 | 1.28 | 2.19 | 0.0002 | 1.50 | 1.23 | 1.84 | <0.0001 | 1.12 | 0.80 | 1.57 | 0.501 | |
| F:SpeechDevelopment | 3.45 | 1.59 | 7.47 | 0.002 | 3.28 | 1.99 | 5.41 | <0.0001 | 1.11 | 0.44 | 2.80 | 0.825 | |
| R:FebrileSeizures | 1.20 | 0.50 | 2.92 | 0.684 | 1.09 | 0.56 | 2.11 | 0.798 | 1.10 | 0.36 | 3.32 | 0.869 | |
| J:Asthma | 1.65 | 1.20 | 2.26 | 0.002 | 1.53 | 1.22 | 1.92 | 0.0003 | 1.08 | 0.73 | 1.59 | 0.696 | |
| L:OtherDermatitisEczema | 1.52 | 1.04 | 2.23 | 0.032 | 1.41 | 1.01 | 1.99 | 0.046 | 1.08 | 0.65 | 1.81 | 0.764 | |
| R:NauseaVomiting | 3.36 | 2.09 | 5.41 | <0.0001 | 3.16 | 2.13 | 4.69 | <0.0001 | 1.07 | 0.58 | 1.98 | 0.836 | |
| P:NeonatalJaundice | 1.40 | 0.99 | 1.97 | 0.058 | 1.32 | 1.01 | 1.72 | 0.039 | 1.06 | 0.69 | 1.64 | 0.780 | |

| Condition | Sex: female | | | | Sex: male | | | | Comparison female vs. male | | | | |
|---|---|---|---|---|---|---|---|---|---|---|---|---|---|
| | $HR_{female}$ | 95%CI LL | 95%CI UL | p-value | $HR_{male}$ | 95%CI LL | 95%CI UL | p-value | $HR_{female}$/ $HR_{male}$ | 95%CI LL | 95%CI UL | p-value | Significant (*) after FDR/BH |
| P:PerinatRespiratCardiovasc | 1.31 | 0.88 | 1.95 | 0.178 | 1.30 | 0.96 | 1.76 | 0.093 | 1.04 | 0.63 | 1.72 | 0.873 | |
| K:Hernia | 1.34 | 0.59 | 3.02 | 0.487 | 1.44 | 1.00 | 2.06 | 0.049 | 1.03 | 0.42 | 2.50 | 0.956 | |
| P:FetalGrowthRetardation | 1.25 | 0.97 | 1.60 | 0.081 | 1.23 | 1.01 | 1.51 | 0.042 | 1.02 | 0.74 | 1.40 | 0.912 | |
| ChapterF:MentalConditions | 3.84 | 2.96 | 4.98 | <0.0001 | 3.90 | 3.25 | 4.67 | <0.0001 | 1.01 | 0.73 | 1.38 | 0.968 | |
| F:OtherDevelopment | 11.38 | 4.35 | 29.77 | <0.0001 | 12.37 | 5.98 | 25.58 | <0.0001 | 1.01 | 0.30 | 3.35 | 0.988 | |
| P:PerinatInfections | 1.80 | 0.99 | 3.25 | 0.053 | 1.79 | 1.09 | 2.94 | 0.020 | 1.01 | 0.48 | 2.14 | 0.976 | |
| Q:NervousSystemMalform | 11.98 | 5.87 | 24.47 | <0.0001 | 10.67 | 5.42 | 21.02 | <0.0001 | 0.98 | 0.36 | 2.67 | 0.970 | |
| ChapterP:PerinatalConditions | 1.20 | 0.95 | 1.51 | 0.120 | 1.25 | 1.05 | 1.50 | 0.014 | 0.97 | 0.72 | 1.29 | 0.813 | |
| J:AcuteLowerRespiratory | 1.55 | 1.12 | 2.13 | 0.008 | 1.62 | 1.26 | 2.08 | 0.0001 | 0.95 | 0.63 | 1.42 | 0.786 | |
| Q:MusculoskeletalMalform | 1.95 | 1.19 | 3.20 | 0.009 | 2.05 | 1.39 | 3.04 | 0.0003 | 0.95 | 0.51 | 1.80 | 0.887 | |
| K:OtherUpperGastrointest | 4.81 | 2.35 | 9.81 | <0.0001 | 5.23 | 2.75 | 9.95 | <0.0001 | 0.87 | 0.33 | 2.28 | 0.776 | |
| P:MaternalComplications | 1.03 | 0.64 | 1.67 | 0.903 | 1.19 | 0.83 | 1.71 | 0.338 | 0.86 | 0.47 | 1.58 | 0.630 | |
| P:PerinatFeeding | 1.86 | 0.93 | 3.73 | 0.079 | 2.21 | 1.20 | 4.06 | 0.011 | 0.85 | 0.34 | 2.13 | 0.725 | |
| E:VolumeDepletion | 2.44 | 1.10 | 5.42 | 0.029 | 2.98 | 1.78 | 4.99 | <0.0001 | 0.81 | 0.31 | 2.10 | 0.666 | |
| P:PerinatHematologic | 1.11 | 0.53 | 2.33 | 0.783 | 1.46 | 0.83 | 2.59 | 0.192 | 0.76 | 0.30 | 1.93 | 0.562 | |
| S:IntracranialInjury | 0.94 | 0.53 | 1.67 | 0.844 | 1.29 | 0.89 | 1.88 | 0.175 | 0.73 | 0.37 | 1.44 | 0.366 | |
| L:Urticaria | 1.15 | 0.57 | 2.32 | 0.687 | 1.62 | 1.00 | 2.62 | 0.049 | 0.71 | 0.30 | 1.66 | 0.432 | |
| K:Stomatitis | 2.29 | 0.95 | 5.52 | 0.064 | 3.44 | 1.86 | 6.34 | <0.0001 | 0.67 | 0.24 | 1.82 | 0.427 | |
| ChapterI:CirculatoryConditions | 1.44 | 0.64 | 3.25 | 0.381 | 2.21 | 1.36 | 3.60 | 0.001 | 0.65 | 0.25 | 1.70 | 0.381 | |
| K:CeliacDisease | 1.71 | 0.81 | 3.63 | 0.161 | 2.62 | 1.34 | 5.13 | 0.005 | 0.64 | 0.23 | 1.76 | 0.383 | |
| G:CerebralPalsy | 4.42 | 2.06 | 9.50 | 0.0001 | 7.27 | 4.36 | 12.12 | <0.0001 | 0.59 | 0.23 | 1.47 | 0.257 | |
| G:Epilepsy | 3.42 | 1.67 | 7.00 | 0.0008 | 7.48 | 4.99 | 11.23 | <0.0001 | 0.46 | 0.20 | 1.06 | 0.069 | |
| R:Dysphagia | 6.18 | 2.66 | 14.35 | <0.0001 | 14.21 | 8.45 | 23.89 | <0.0001 | 0.43 | 0.16 | 1.15 | 0.093 | |
| F:OtherAnxietyConditions | 1.46 | 0.69 | 3.09 | 0.325 | 3.70 | 2.11 | 6.50 | <0.0001 | 0.38 | 0.15 | 0.97 | 0.043 | |

Sex-specific hazard ratios in female individuals with ARFID vs. female controls ($HR_{female}$) and male individuals with ARFID vs. male controls ($HR_{male}$), lower/upper limits of the cluster-robust 95% confidence interval (95% CI LL/UL), and raw p-values are stated for each analyzed condition (conditions with n<5 individuals per sex in ARFID or controls were not analyzed) from the sex-stratified Cox regression model (predictor: group [ARFID vs. controls]; sex strata [female/male strata]; covariate: birth year [1992–2008, factorized]; robust sandwich estimates given clustered twin data). Sex-specific HRs were compared ($HR_{female}$/$HR_{male}$) and statistics of the comparison are stated: lower/upper limits of the cluster-robust 95% confidence interval of $HR_{female}$/$HR_{male}$, raw p-value, and significance according to the false discovery rate-adjusted threshold (FDR/Benjamini-Hochberg procedure [BH]). There were no statistically significant sex differences ($HR_{female}$ vs. $HR_{male}$) in ARFID vs. controls at $\alpha_{FDR}$=0.00014 (across 103 supplementary tests). Nominally significant sex differences (p<0.050) are highlighted with light-orange background. Conditions are sorted in descending order of $HR_{female}$/$HR_{male}$ for easier orientation. *Abbreviations: ADHD, attention deficit hyperactivity disorder; ARFID, avoidant restrictive food intake disorder; CNS, central nervous system; GERD, gastro-esophageal reflux disorder; IBD, inflammatory bowel disease.*

**Table S9. Sex effects on number of distinct overall, mental, and somatic diagnoses in ARFID vs. controls**

| Condition | ARFID | | | | Controls | | | | Poisson regression model statistics | | | | | | |
|---|---|---|---|---|---|---|---|---|---|---|---|---|---|---|---|
| | Female | | Male | | Female | | Male | | Group-*x*-sex interaction effect | | | | | | |
| | **Mean** | **SEM** | **Mean** | **SEM** | **Mean** | **SEM** | **Mean** | **SEM** | **IRR[a]** | **95%CI LL** | **95%CI UL** | **Robust SE** | **z statistic** | **p-value** | **Significant (*) after FDR/BH** |
| All diagnoses | 5.85 | 0.44 | 6.53 | 0.37 | 2.54 | 0.03 | 3.01 | 0.03 | 1.05 | 0.87 | 1.27 | 0.10 | 0.55 | 0.580 | |
| Mental diagnoses | 0.77 | 0.10 | 1.16 | 0.10 | 0.19 | 0.01 | 0.25 | 0.01 | 0.85 | 0.61 | 1.18 | 0.17 | -0.96 | 0.335 | |
| Somatic diagnoses | 5.08 | 0.41 | 5.38 | 0.32 | 2.36 | 0.02 | 2.77 | 0.03 | 1.11 | 0.90 | 1.35 | 0.10 | 0.97 | 0.330 | |

Mean and standard error of the mean (SEM) for number of *all* distinct diagnoses (i.e., unique diagnostic ICD-codes), number of distinct *mental* diagnoses, and number of distinct *somatic* diagnoses per individual twin are given in female and male individuals with ARFID and female and male controls. As test statistics, incidence rate ratio of the group (ARFID vs. controls)-*x*-sex interaction effect (IRR), lower/upper limits of the cluster-robust 95% confidence interval (95% CI LL/UL), cluster-robust standard error (SE), z statistic, raw p-value, and significance according to supplementary false discovery rate-adjusted threshold (FDR/Benjamini-Hochberg procedure [BH]) are stated for all distinct diagnoses, distinct mental diagnoses, and distinct somatic diagnoses. Statistics were obtained with Poisson regression models including a group-*x*-sex interaction term (other predictors/main effects: group [ARFID vs. controls], sex [female/male]; covariate: birth year [1992–2008, factorized]; robust sandwich estimates given clustered twin data; offset term to account for follow-up/exposure time). There were no significant group-*x*-sex interaction effects at $\alpha_{FDR}$=0.00014 (across 103 supplementary tests). [a]IRRs>1 indicate greater incidence rates in females with ARFID than males with ARFID. IRRs<1 indicate greater incidence rates in males with ARFID than females with ARFID (regarding directionality [*not* statistical significance]). *Abbreviation: ARFID, avoidant restrictive food intake disorder.*

**Table S10. Sex effects on number of inpatient days due to any, any mental, and any somatic diagnosis in ARFID vs. controls**

| Condition | ARFID | | | | Controls | | | | Poisson regression model statistics | | | | | | |
|---|---|---|---|---|---|---|---|---|---|---|---|---|---|---|---|
| | Female | | Male | | Female | | Male | | Group-*x*-sex interaction effect | | | | | | |
| | Mean | SEM | Mean | SEM | Mean | SEM | Mean | SEM | IRR[a] | 95%CI LL | 95%CI UL | Robust SE | z statistic | p-value | Significant (*) after FDR/BH |
| Any diagnosis | 29.57 | 5.61 | 24.71 | 2.65 | 8.82 | 0.19 | 9.54 | 0.19 | 1.28 | 0.82 | 1.97 | 0.22 | 1.09 | 0.274 | |
| Mental diagnosis | 2.05 | 1.48 | 0.32 | 0.12 | 0.39 | 0.07 | 0.09 | 0.02 | 1.35 | 0.25 | 7.32 | 0.86 | 0.34 | 0.730 | |
| Somatic diagnosis | 25.44 | 5.07 | 21.48 | 2.34 | 8.00 | 0.16 | 8.96 | 0.18 | 1.31 | 0.83 | 2.06 | 0.23 | 1.17 | 0.244 | |

Mean and standard error of the mean (SEM) for number of inpatient days per individual twin due to any diagnosis (i.e., unique diagnostic ICD-code), any mental diagnosis (as main diagnosis/cause of hospitalization), and any somatic diagnosis (as main diagnosis/cause of hospitalization) are given in female and male individuals with ARFID and female and male controls. As test statistics, incidence rate ratio of the group (ARFID vs. controls)-*x*-sex interaction effect (IRR), lower/upper limits of the cluster-robust 95% confidence interval (95% CI LL/UL), cluster-robust standard error (SE), z statistic, raw p-value, and significance according to supplementary false discovery rate-adjusted threshold (FDR/Benjamini-Hochberg procedure [BH]) are stated for any diagnosis, any mental diagnosis, and any somatic diagnosis. Statistics were obtained with Poisson regression models including a group-*x*-sex interaction term (other predictors/main effects: group [ARFID vs. controls], sex [female/male]; covariate: birth year [1992–2008, factorized]; robust sandwich estimates given clustered twin data; offset term to account for follow-up/exposure time). There were no significant group-*x*-sex interaction effects at $\alpha_{FDR}$=0.00014 (across 103 supplementary tests). [a]IRRs>1 indicate greater incidence rates in females with ARFID than males with ARFID. IRRs<1 indicate greater incidence rates in males with ARFID than females with ARFID (regarding directionality [*not* statistical significance]). *Abbreviation: ARFID, avoidant restrictive food intake disorder.*

**Table S11. Age group-stratified Cox regression model estimates in ARFID vs. controls**

| Condition | Age group 1: 0 to <6 years | | | | Age group 2: 6 to <12 years | | | | Age group 3: 12 to <18 years | | | | Comparison age groups 1 vs. 2 | | | | | Comparison age groups 3 vs. 2 | | | | |
|---|---|---|---|---|---|---|---|---|---|---|---|---|---|---|---|---|---|---|---|---|---|---|
| | $HR_{age\ group\ 1}$ | 95%CI LL | 95%CI UL | p-value | $HR_{age\ group\ 2}$ | 95%CI LL | 95%CI UL | p-value | $HR_{age\ group\ 3}$ | 95%CI LL | 95%CI UL | p-value | $HR_{age\ group\ 1}$/ $HR_{age\ group\ 2}$ | 95%CI LL | 95%CI UL | p-value | Sig. (*) after FDR | $HR_{age\ group\ 3}$/ $HR_{age\ group\ 2}$ | 95%CI LL | 95%CI UL | p-value | Sig. (*) after FDR |
| F:Autism | 16.09 | 9.40 | 27.56 | <0.0001 | 12.35 | 8.75 | 17.43 | <0.0001 | 4.32 | 2.54 | 7.34 | <0.0001 | 1.30 | 0.69 | 2.46 | 0.413 | | 0.35 | 0.19 | 0.66 | 0.001 | |
| ChapterF:MentalConditions | 5.17 | 3.95 | 6.77 | <0.0001 | 4.48 | 3.67 | 5.47 | <0.0001 | 2.12 | 1.52 | 2.95 | <0.0001 | 1.15 | 0.83 | 1.61 | 0.399 | | 0.47 | 0.32 | 0.70 | 0.0001 | * |
| ChapterI:CirculatoryConditions | 2.19 | 1.12 | 4.30 | 0.023 | 2.50 | 1.36 | 4.59 | 0.003 | 1.17 | 0.48 | 2.83 | 0.733 | 0.88 | 0.35 | 2.18 | 0.777 | | 0.47 | 0.17 | 1.28 | 0.140 | |
| ChapterE:EndocrineConditions | 3.10 | 2.32 | 4.15 | <0.0001 | 2.97 | 2.10 | 4.21 | <0.0001 | 1.53 | 0.89 | 2.65 | 0.126 | 1.04 | 0.66 | 1.64 | 0.858 | | 0.52 | 0.27 | 0.98 | 0.044 | |
| F:ADHD | 9.14 | 4.33 | 19.32 | <0.0001 | 5.87 | 4.51 | 7.65 | <0.0001 | 3.10 | 2.08 | 4.60 | <0.0001 | 1.56 | 0.70 | 3.44 | 0.274 | | 0.53 | 0.33 | 0.85 | 0.009 | |
| Q:MusculoskeletalMalform | 1.70 | 1.12 | 2.57 | 0.013 | 3.37 | 1.92 | 5.93 | <0.0001 | 1.79 | 0.79 | 4.04 | 0.16 | 0.50 | 0.25 | 1.01 | 0.055 | | 0.53 | 0.20 | 1.43 | 0.211 | |
| F:IntellectualDisability | 16.18 | 9.62 | 27.21 | <0.0001 | 9.82 | 6.29 | 15.33 | <0.0001 | 5.98 | 2.82 | 12.66 | <0.0001 | 1.65 | 0.83 | 3.26 | 0.152 | | 0.61 | 0.25 | 1.46 | 0.265 | |
| S:IntracranialInjury | 1.45 | 0.94 | 2.25 | 0.092 | 1.12 | 0.66 | 1.91 | 0.667 | 0.74 | 0.33 | 1.65 | 0.461 | 1.29 | 0.65 | 2.57 | 0.461 | | 0.66 | 0.25 | 1.73 | 0.394 | |
| K:NoninfectiveIBD | 2.02 | 1.21 | 3.38 | 0.008 | 4.42 | 2.18 | 8.97 | <0.0001 | 2.96 | 1.19 | 7.39 | 0.02 | 0.46 | 0.19 | 1.10 | 0.081 | | 0.67 | 0.21 | 2.12 | 0.495 | |
| J:Asthma | 1.54 | 1.23 | 1.92 | 0.0001 | 1.77 | 1.22 | 2.57 | 0.002 | 1.22 | 0.61 | 2.44 | 0.584 | 0.87 | 0.57 | 1.33 | 0.514 | | 0.69 | 0.31 | 1.51 | 0.350 | |
| ChapterK:DigestiveConditions | 1.95 | 1.61 | 2.35 | <0.0001 | 2.33 | 1.79 | 3.03 | <0.0001 | 1.77 | 1.19 | 2.64 | 0.005 | 0.84 | 0.61 | 1.15 | 0.267 | | 0.76 | 0.47 | 1.23 | 0.263 | |
| J:ChronicUpperRespiratory | 1.41 | 1.08 | 1.85 | 0.013 | 1.21 | 0.83 | 1.78 | 0.322 | 0.97 | 0.51 | 1.87 | 0.937 | 1.16 | 0.74 | 1.84 | 0.516 | | 0.80 | 0.38 | 1.72 | 0.572 | |
| ChapterJ:RespiratoryConditions | 1.43 | 1.25 | 1.63 | <0.0001 | 1.39 | 1.04 | 1.85 | 0.028 | 1.30 | 0.84 | 2.02 | 0.238 | 1.03 | 0.75 | 1.42 | 0.847 | | 0.94 | 0.55 | 1.59 | 0.817 | |
| J:AcuteUpperRespiratory | 1.38 | 1.16 | 1.64 | 0.0002 | 1.80 | 1.21 | 2.69 | 0.004 | 1.75 | 0.93 | 3.26 | 0.081 | 0.76 | 0.50 | 1.18 | 0.225 | | 0.97 | 0.46 | 2.03 | 0.930 | |
| J:Rhinitis | 1.25 | 0.62 | 2.54 | 0.53 | 1.57 | 1.08 | 2.29 | 0.017 | 1.56 | 0.91 | 2.68 | 0.107 | 0.80 | 0.36 | 1.77 | 0.577 | | 0.99 | 0.51 | 1.91 | 0.979 | |
| K:Constipation | 1.93 | 1.39 | 2.69 | <0.0001 | 2.50 | 1.75 | 3.56 | <0.0001 | 2.56 | 1.36 | 4.85 | 0.004 | 0.77 | 0.47 | 1.26 | 0.303 | | 1.03 | 0.50 | 2.13 | 0.944 | |
| AllergicConditions | 1.52 | 1.24 | 1.87 | <0.0001 | 1.59 | 1.17 | 2.15 | 0.003 | 1.65 | 1.06 | 2.57 | 0.027 | 0.96 | 0.67 | 1.38 | 0.815 | | 1.04 | 0.61 | 1.76 | 0.892 | |
| ChapterQ:CongenitalConditions | 1.96 | 1.60 | 2.40 | <0.0001 | 1.43 | 0.88 | 2.32 | 0.154 | 2.15 | 1.24 | 3.72 | 0.006 | 1.38 | 0.81 | 2.33 | 0.234 | | 1.51 | 0.72 | 3.14 | 0.274 | |
| R:Dysphagia | 11.84 | 6.34 | 22.12 | <0.0001 | 7.25 | 3.27 | 16.08 | <0.0001 | 13.86 | 5.85 | 32.81 | <0.0001 | 1.63 | 0.59 | 4.49 | 0.342 | | 1.91 | 0.59 | 6.16 | 0.278 | |
| AutoimmuneConditions | 1.36 | 0.95 | 1.95 | 0.093 | 0.90 | 0.51 | 1.59 | 0.717 | 1.77 | 0.99 | 3.15 | 0.053 | 1.51 | 0.78 | 2.95 | 0.223 | | 1.97 | 0.87 | 4.43 | 0.103 | |
| L:OtherDermatitisEczema | 1.53 | 1.10 | 2.12 | 0.012 | 0.86 | 0.49 | 1.53 | 0.616 | 2.45 | 1.47 | 4.10 | 0.0006 | 1.77 | 0.91 | 3.43 | 0.093 | | 2.84 | 1.32 | 6.11 | 0.008 | |

Age group-specific hazard ratios in 0 to <6-year-old individuals with ARFID vs. controls of the same age group ($HR_{age\ group\ 1}$), 6 to <12-year-old individuals with ARFID vs. controls ($HR_{age\ group\ 2}$), and 12 to <18-year-old individuals with ARFID vs. controls ($HR_{age\ group\ 3}$), lower/upper limits of the cluster-robust 95% confidence interval (95% CI LL/UL), and p-values are stated for each analyzed condition (conditions with n<5 individuals per age group in ARFID or controls were not analyzed) from the age group-stratified Cox regression model (predictor: group [ARFID vs. controls]; age/time group strata [age groups 1-3]; covariates: sex [female/male], birth year [1992–2008, factorized]; robust sandwich estimates given clustered twin data). Age group-specific HRs were compared ($HR_{age\ group\ 1}$/$HR_{age\ group\ 2}$ and $HR_{age\ group\ 3}$/$HR_{age\ group\ 2}$) and statistics of both comparisons are stated: lower/upper limits of the cluster-robust 95% confidence interval of $HR_{age\ group\ 1}$/$HR_{age\ group\ 2}$ and $HR_{age\ group\ 3}$/$HR_{age\ group\ 2}$, raw p-value, and significance according to the false discovery rate-adjusted threshold (FDR/Benjamini-Hochberg procedure [BH]). There was one significant age group difference ($HR_{age\ group\ 3}$ vs. $HR_{age\ group\ 2}$ for ChapterF:MentalConditions, marked by an asterisk [*] and orange background) in ARFID vs. controls at $\alpha_{FDR}$=0.00014 (across 103 supplementary tests). All other nominally significant age group differences (p<0.050) are highlighted with light-orange background. Conditions are sorted in ascending order of $HR_{age\ group\ 3}$/$HR_{age\ group\ 2}$ for easier orientation. *Abbreviations: ADHD, attention deficit hyperactivity disorder; ARFID, avoidant restrictive food intake disorder; IBD, inflammatory bowel disease.*

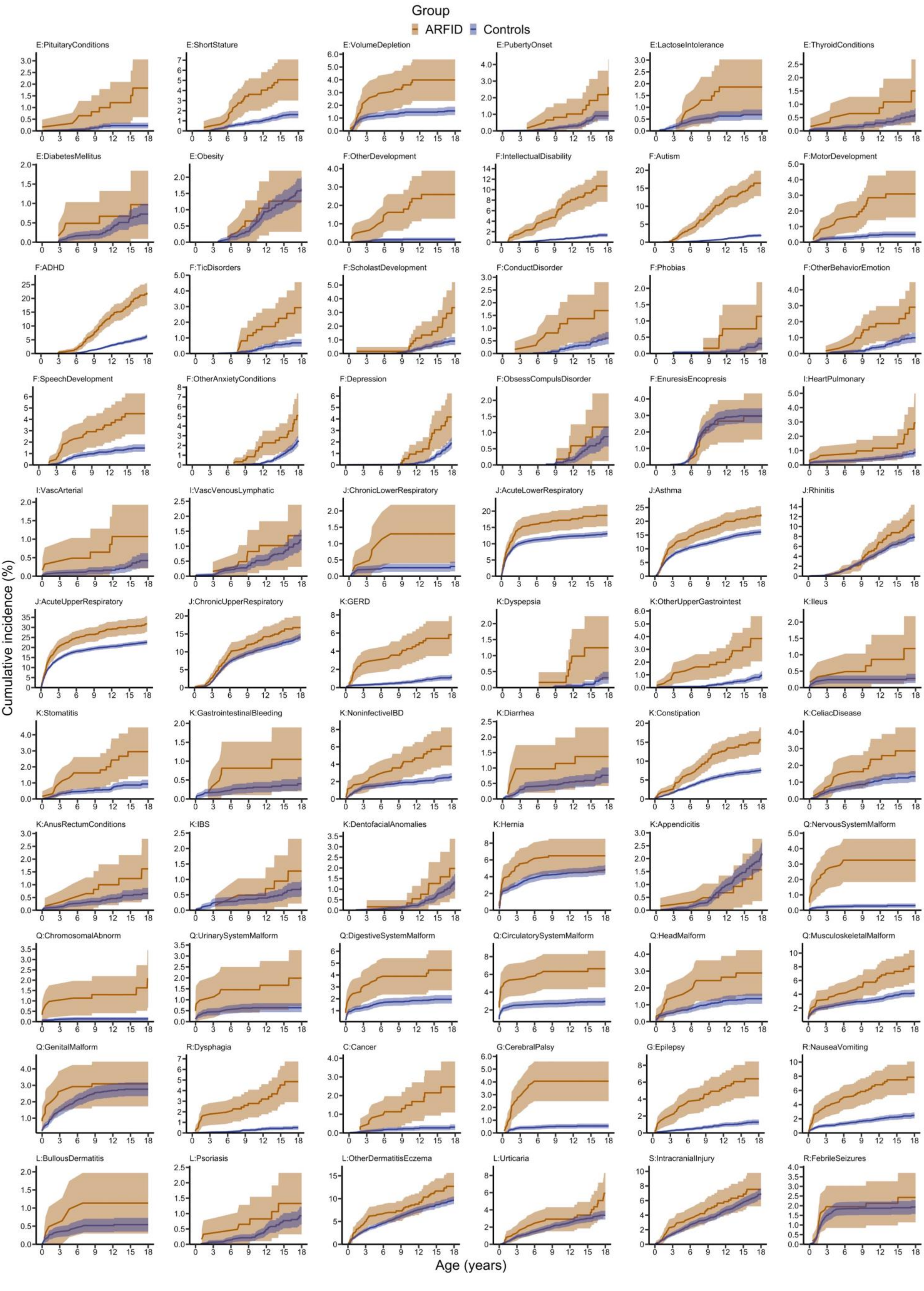
Group
ARFID
Controls
E:PituitaryConditions
E:ShortStature
E:VolumeDepletion
E:PubertyOnset
E:LactoseIntolerance
E:ThyroidConditions
E:DiabetesMellitus
E:Obesity
F:OtherDevelopment
F:IntellectualDisability
F:Autism
F:MotorDevelopment
F:ADHD
F:TicDisorders
F:ScholastDevelopment
F:ConductDisorder
F:Phobias
F:OtherBehaviorEmotion
F:SpeechDevelopment
F:OtherAnxietyConditions
F:Depression
F:ObsessCompulsDisorder
F:EnuresisEncopresis
I:HeartPulmonary
I:VascArterial
I:VascVenousLymphatic
J:ChronicLowerRespiratory
J:AcuteLowerRespiratory
J:Asthma
J:Rhinitis
J:AcuteUpperRespiratory
J:ChronicUpperRespiratory
K:GERD
K:Dyspepsia
K:OtherUpperGastrointest
K:Ileus
K:Stomatitis
K:GastrointestinalBleeding
K:NoninfectiveIBD
K:Diarrhea
K:Constipation
K:CeliacDisease
K:AnusRectumConditions
K:IBS
K:DentofacialAnomalies
K:Hernia
K:Appendicitis
Q:NervousSystemMalform
Q:ChromosomalAbnorm
Q:UrinarySystemMalform
Q:DigestiveSystemMalform
Q:CirculatorySystemMalform
Q:HeadMalform
Q:MusculoskeletalMalform
Q:GenitalMalform
R:Dysphagia
C:Cancer
G:CerebralPalsy
G:Epilepsy
R:NauseaVomiting
L:BullousDermatitis
L:Psoriasis
L:OtherDermatitisEczema
L:Urticaria
S:IntracranialInjury
R:FebrileSeizures
Cumulative incidence (%)
Age (years)

**Figure S1. Cumulative incidence plots in ARFID vs. controls for individual and grouped conditions**

Cumulative incidence of having a *specific* condition (%) and lower and upper limits of its cluster-robust 95% confidence interval are plotted over age (0 to <18 years) in ARFID vs. controls. The Kaplan-Meier-based estimation method is explained in the statistical analysis section of the main article and in the legend of Table S5. Conditions are sorted in accordance with Table S4, i.e., in descending order of base Cox regression model hazard ratios within ICD-chapters, the group of further conditions, and grouped allergic and autoimmune conditions (please consult Figure 4 in the main article for cumulative incidence plots for ICD-*chapters* and grouped allergic and autoimmune conditions; perinatal conditions excluded given their occurrence during the perinatal period and lack of age-related dynamics). *Abbreviations: ADHD, attention deficit hyperactivity disorder; ARFID, avoidant restrictive food intake disorder; GERD, gastro-esophageal reflux disorder; IBD, inflammatory bowel disease; IBS, irritable bowel syndrome.*

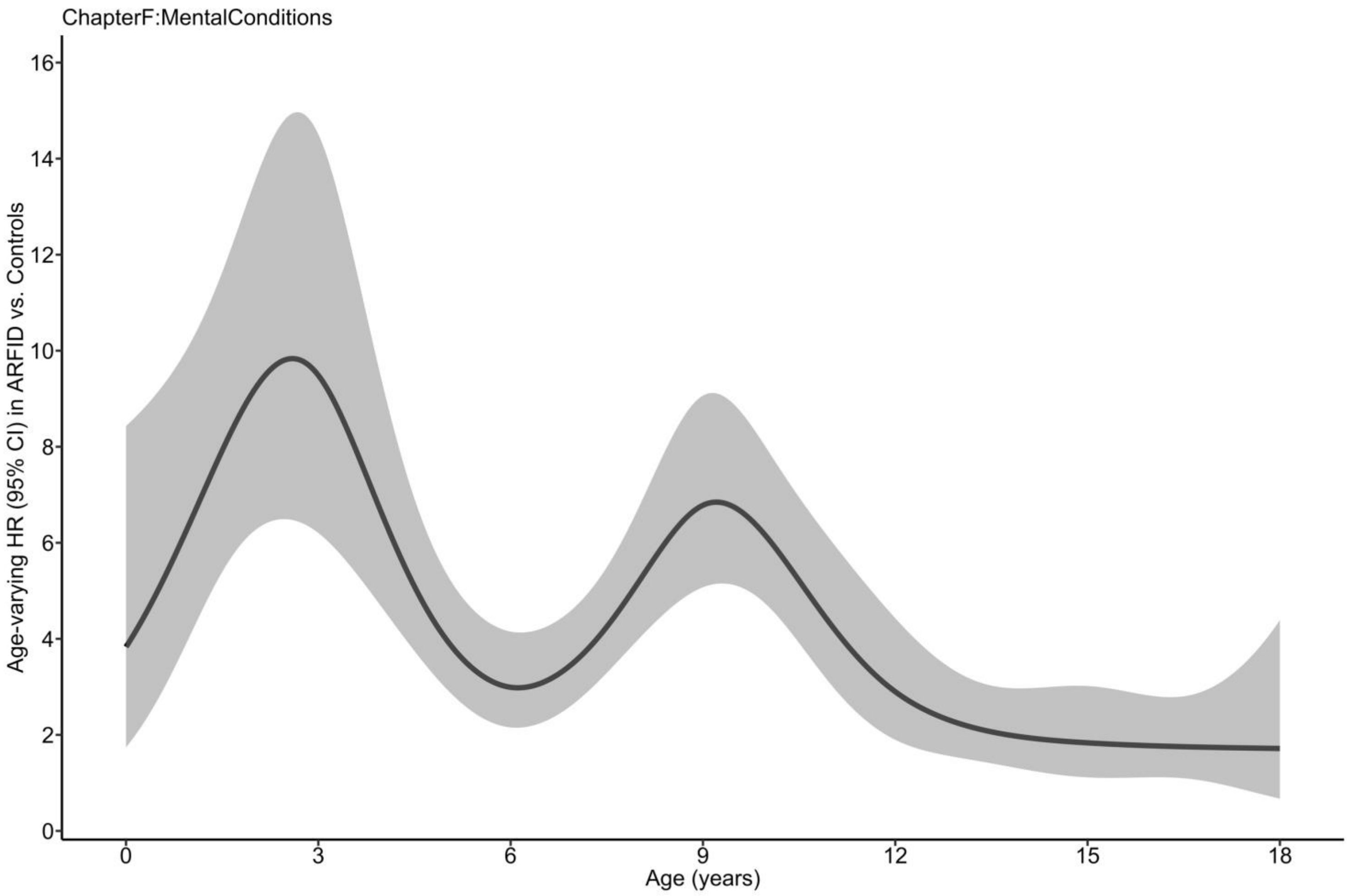


**Figure S2. Age-varying hazard ratios in ARFID vs. controls for ICD-chapter F: mental conditions**

Following-up on significant age group differences ($HR_{age\ group\ 3}$ vs. $HR_{age\ group\ 2}$) in ARFID vs. controls for ICD-chapter F: mental conditions (Table S11), time-varying hazard ratios (HRs) in ARFID vs. controls and lower and upper limits of their cluster-robust 95% confidence intervals are plotted over age (0 to <18 years). The curve was estimated using the base Cox regression model (predictor: group [ARFID vs. controls]; covariates: sex [female/male], birth year [1992–2008, factorized]; robust sandwich estimates given clustered twin data) and adding age-/time-related variance via cubic splines with pre-specified knots at 3, 6, 9, 12, and 15 years of age (boundary knots at 0 and <18 [≈17.9] years of age). *Abbreviation: ARFID, avoidant restrictive food intake disorder.*
